# Pay-it-forward to improve influenza vaccine uptake and public engagement among children and older adults in China: A quasi-experimental pragmatic trial

**DOI:** 10.1101/2021.10.30.21265713

**Authors:** Dan Wu, Chenqi Jin, Khaoula Bessame, Fanny Fong-Yi Tang, Jason J. Ong, Zaisheng Wang, Yewei Xie, Mark Jit, Heidi J. Larson, Tracey Chantler, Leesa Lin, Wenfeng Gong, Fan Yang, Fengshi Jing, Shufang Wei, Weibin Cheng, Yi Zhou, Nina Ren, Shuhao Qiu, Jianmin Bao, Liufen Wen, Qinlu Yang, Junzhang Tian, Weiming Tang, Joseph D. Tucker

## Abstract

**Background:** China has low seasonal influenza vaccination rates among children and older adults. This quasi-experimental pragmatic trial examined the effectiveness of a pay-it-forward intervention on influenza vaccination compared to standard-of-care (user-paid vaccination) among children and older people in China. Pay-it-forward offered a free influenza vaccine from a local group and an opportunity to donate financially to support future individuals.

**Methods:** At each of the three study sites, participants were recruited into the standard-of-care arm first until expected sample size was reached and then recruited into the pay-it-forward arm. The primary outcome was vaccine uptake. Secondary outcomes included vaccine confidence and costs. Regression methods compared influenza vaccine uptake and vaccine confidence between the two arms.

**Results:** Among 300 participants enrolled, 55/150(36.7%) in the standard-of-care arm 111/150(74.0%) in the pay-it-forward arm received an influenza vaccine. People in the pay-it-forward arm were more likely to receive the vaccine compared to people in the standard-of-care arm (adjusted odds ratio (aOR)= 6.7, 95%CI [2.7, 16.6] among children; aOR=5.0, [2.3, 10.8] among older adults). People in the pay-it-forward arm had greater confidence in vaccine safety, importance, and effectiveness. In the pay-it-forward arm, 107/111 (96.4%) of participants donated money for subsequent vaccinations, and 19 of 60 invited (31.7%) created postcard messages. The pay-it-forward arm had a lower economic cost per person vaccinated ($45.60) than the standard-of-care arm ($64.67).

**Conclusions:** Pay-it-forward was effective in improving influenza vaccine uptake and public engagement. Our data have implications for pro-social interventions to enhance influenza vaccine uptake in countries where influenza vaccines are available for a fee.

**Trial registration:** ChiCTR2000040048

**Main point summary:** Pay-it-forward substantially increased influenza vaccine uptake among children and older adults compared to standard of care user-paid vaccination.

## BACKGROUND

In mainland China, an average of 10 people die from influenza-related illnesses each hour.^1^ Influenza vaccination is the most effective way to prevent morbidity and mortality attributable to influenza.^2^ Influenza vaccine is increasingly important during COVID-19 because it might help reduce risks of acquiring SARS-CoV-2 that causes COVID-19.^3,4^ The Chinese Center for Disease Control and Prevention (China CDC) guidelines recommend influenza vaccination for high-risk populations, including children and older adults. However, influenza immunization policies widely vary^5^, and most cities in China do not provide free influenza vaccines to high-risk individuals. A meta-analysis reported less than one-fifth of children and older adults in China received an influenza vaccine in the past year.^6^ Low influenza vaccine uptake is common in many other low- and middle-income countries.^7^

There are several reasons for low influenza vaccination uptake in China.^6,8^ First, most people in China are unaware of influenza vaccination, and many people are hesitant about vaccine safety and effectiveness.^9^ Second, there is minimal public engagement in vaccinations.^10^ Despite a strong rationale for public engagement, few programs engage the public regarding influenza vaccinations. Third, there are limited public resources to support influenza vaccination among high-risk populations. The influenza vaccine is largely not covered by mandatory health insurance schemes, and, as a result, most people have to pay US$8.5-23.5 out-of-pocket to be vaccinated.^11^ Innovative strategies are needed to improve influenza vaccine uptake.

Pay-it-forward is a community-engaged social innovation, which has an individual receive a free influenza vaccine and a handwritten postcard message co-created by previous participants informing them that someone else has paid for them to receive a free vaccine.^12^ After receiving the vaccination, the recipients are asked if they would like to support the vaccination of a subsequent person (supplementary Fig 1). Our previous pay-it-forward studies focused on increasing testing for sexually transmitted infections among sexual minorities in gay men led sexual health clinics. The pay-it-forward arm had a chlamydia and gonorrhea dual test uptake of 56% compared to 18% in the standard-of-care arm, where participants had to pay out-of-pocket.^13,14^ Over 90% of participants donated to the rolling finance pool, and qualitative data showed that trust in health services improved among participants in the pay-it-forward arm.^15^ But pay-it-forward has not been examined in increasing vaccination services uptake in community-based primary care facilities in the public sector.

This quasi-experimental pragmatic trial assessed the effectiveness of a pay-it-forward intervention to increase influenza vaccination uptake at three study sites among children (aged between 6 months and eight years) and older adults (aged 60 or above) in comparison to the current standard-of-care (user-paid vaccination) in China.

## METHODS

### Study design and participants

Guangdong is a subtropical province in southern China with over 120 million people. In Southern China, influenza is prevalent throughout the year.^16^ In this study, we selected three study sites where influenza vaccination was available on a fee basis. These three study sites were: a rural site (Yangshan), a suburban site (Zengcheng), and an urban site (Tianhe). Clinics were selected because they had sufficient influenza vaccines in stock and medical staff (nurses, doctors) familiar with influenza vaccination.

This study consisted of three stages 1) co-creation of the intervention and engagement strategies with stakeholders during a three-day hackathon; 2) a feasibility pilot to inform the recruitment process and sample size calculations; and 3) a pay-it-forward quasi-experimental pragmatic trial to evaluate the effectiveness of the intervention.

### Public engagement

Three team members joined a participatory hackathon from November 4-6, 2019, to co-create the pay-it-forward intervention. Co-creation is an iterative, bidirectional partnership between researchers and the public to develop new ideas.^17^ Participants included potential end users, public health practitioners, health innovators, communication experts and vaccine experts. We mapped out the following elements of the study: 1) key stakeholders of the study; 2) potential user journeys; 3) engagement strategies; 4) behavioral mechanisms; and 5) donation strategies.

Community engagement strategies used in this study included the following: inviting community members to design postcards relevant to influenza vaccination (Supplementary Fig 2); working in partnership with a local older adult to co-develop a video to explain pay-it-forward (supplementary video link); inviting study participants to write postcard messages during recruitment for future participants (Supplementary Fig 3); and engaging local medical staff in implementing the quasi-experimental study, including having one-to-two staff members at each study site to help adjust recruitment and communication efforts to the local dialect. The community’s six postcard designs were subsequently used to explain the pay-it-forward system to potential participants.

### Pilot

We carried out a feasibility pilot at the rural study site from January to April 2020, during COVID-19 restrictions. The purpose of the pilot was to finalize the pay-it-forward intervention process and estimate effect size based on the primary outcome of vaccine uptake to inform power calculations. This pilot demonstrated that, in the pay-it-forward arm, 90.9% (40/44) of participants received an influenza vaccine. Thirteen of 57 participants (22.3%) in the standard-of-care arm received a vaccine.

### Sample size calculation

Given the differences between children and older adults, we stratified sample size calculations by age groups. Based on our pilot data, we optimistically estimated the vaccine uptake as 30% in the standard-of-care arm, and conservatively estimated the vaccine uptake in the pay-it-forward arm as 80%, and a significance level of 0.025. Thus, a sample size of 100 (50 in the control arm and 50 in the intervention) for each age group would give us 90% power to test that the pay-it-forward is superior to the stand of care in promoting vaccination uptake, with a margin of 10%. We increased the sample size by 50% to allow for secondary analyses, resulting in a sample size of 75 for each age group in each arm.

In addition to the primary comparison (pay-it-forward compared to standard of care), we implemented an exploratory arm (n = 150) that provided free vaccines without any community engagement. We included a free vaccine arm because this provided an opportunity to compare the cost of pay-it-forward and free service provision. However, the study was not powered to assess the difference between pay-it-forward and free arms, and as a result, we include this data as supplementary material.

### Quasi-experimental pragmatic trial

This trial was implemented in rural, suburban, and urban study sites. Each study site implemented all study arms. Because of practical considerations, recruited participants were chronologically allocated (non-random) into the specified study arms (Supplementary Fig 4: time-based recruitment). At each site, the standard-of-care arm was followed by the pay-it-forward. Influenza vaccine services are usually available in China from September to April. Influenza vaccine availability is idiosyncratic at specific health facilities because of the supply chain problems in local settings. Despite discussions with health authorities and vaccine manufacturers, study sites encountered lapses in supply. The time needed to recruit each study arm was related to the availability of vaccines and the number of people willing to participate.

The inclusion criteria for this study differed by age group and were determined according to China’s national influenza vaccine guidelines.^18^ Eligibility criteria were: aged between six months and eight years old (children) or ≥ 60 years old (older adults); no acute moderate or severe illnesses; eligible to receive an influenza vaccine based on clinical evaluation from a physician; has a legal guardian (children) or capable of making informed decisions (older adults); consents to participate in the study; and has not received an influenza vaccine in the past year. If multiple people in a family were eligible to join the study, we allowed one person to join. All eligible children and older adults presenting to these sites were invited to participate by medical staff involved in the study during the recruitment periods.

### Procedures

We were informed of vaccine availability by local study sites and commenced data collection on 21 September 2020. Among all potential participants visiting the selected clinics, medical staff assessed eligibility for the study based on inclusion criteria and introduced eligible participants to project staff.

#### Standard-of-care

Participants recruited in the standard-of-care arm were provided with a brief introduction to the influenza vaccine by project staff using a pamphlet about influenza and influenza vaccination (Supplementary Fig 5). They were then asked if they were willing to pay out of pocket at the standard market price (US$8.5-23.5 depending on the market price of vaccines provided at the clinic) to receive an influenza vaccination. Those who agreed to pay were screened for vaccination eligibility, and those without any contraindications received the vaccine.

#### Pay-it-forward

Participants recruited in the pay-it-forward arm were provided with the same introductory pamphlet about influenza and influenza vaccination. Project staff then explained the pay-it-forward program (Supplementary Fig 6), including its purpose, the opportunity to receive one dose of influenza vaccination for free, and the opportunity to donate money towards someone else’s vaccine dose and write postcard messages. Participants were told that the normal price to receive an influenza vaccine, including administration fees were RMB 56(US$ 8.5) for children and 153(US$ 23.5) for adults, and that previous participants had donated money to cover the costs and had also created handwritten postcard messages for them.

If the participants decided to receive vaccination, they were asked prior to receiving the vaccination whether they were willing to donate any amount of money into a pool of funds to support subsequent participants in receiving the same vaccine. They were assured that the donation was entirely voluntary, and any donation amount was acceptable and would not affect whether they received a vaccination or subsequent care. They were also invited to write anonymous postcard messages for future participants. A donation collection box was provided onsite for those who preferred to donate cash. A QR code using WeChat (a multifunctional social mobile app embedded with anonymous money transfer functions) was provided to those who chose to make online donations.

Donations were used to support the vaccination of subsequent participants and aggregated data on donation amounts were made publicly available on the website and WeChat newsletter of Social Entrepreneurship to Spur Health (a research hub in the UNICEF/UNDP/World Bank/WHO Special Programme for Research and Training in Tropical Diseases Social Innovation in Health Initiative). COVID-19 conditions prevented participants from creating handwritten postcards during some periods of the trial.

Participation was voluntary and anonymous. After introducing the intervention, all participants were asked to complete a short, self-administered online questionnaire to collect information about sociodemographic characteristics and attitudes towards influenza vaccines (supplementary questionnaire). Vaccine confidence in importance, safety, and effectiveness were measured using survey items adapted to assess influenza vaccine confidence in China.^19,20^ Those who had difficulty reading the questionnaire were assisted by the project and healthcare staff onsite. A small gift worth around RMB10 (US$1.5) was given to each participant after completing the questionnaire survey.

Data collection was conducted from September 2020 to March 2021. The study collected the following information: administrative data recorded by research staff using a standard information tracking sheet including the number of invited and participating individuals, the number of participants who received the vaccine, as well as survey data through a self-administered survey instrument. Administrative and survey data were linked using numerical IDs. We collected information about the number of participants in the pay-it-forward arm who donated and corresponding donation amount, and those who created a postcard text for subsequent people. Economic costs were collected.

### Study outcomes

The primary outcome was influenza vaccine uptake ascertained by administrative records. Secondary outcomes included self-reported vaccine confidence, defined as public trust in the vaccine safety, importance and effectiveness ^21^ and economic evaluation.

### Data analysis

Descriptive analyses were conducted to summarize sociodemographic and behavioral characteristics, participation rate, and vaccination rate. We used a Chi-squared test to investigate differences in vaccination uptake between the standard-of-care and pay-it-forward arms. We ran multivariable logistic regression to examine the association between outcomes and interventions (standard-of-care and pay-it-forward) after adjusting for potential confounders. We summarized the participants’ donations in the pay-it-forward arm and compared proportions of participants between rural, suburban, and urban sites who contributed US$7.6 (close to a child vaccine cost) or more. All data were analyzed using SPSS Version 25 and STATA 17.

### Cost Analysis

We evaluated the costs of the standard-of-care and pay-it-forward arms using a micro-costing approach and reported this in 2020 USD. The costs of implementing each arm were estimated using invoices, onsite staff’s self-reporting the wages of healthcare workers, and estimated opportunity costs of community staff’s time (supplementary costs file). Additional costs in the pay-it-forward arm related to volunteer time and the recruitment and donation process associated with the start-up and recurrent costs. Financial costs were obtained by subtracting donation contributions from the total economic cost. The analysis was performed from the healthcare provider’s perspective, the Guangdong Department of Health. We reported the total economic and financial cost for each arm and the cost per person vaccinated.

## RESULTS

In total, 174 children’s caregivers and 182 older adults were approached at the three study sites (Fig 1). Forty-one people declined to participate, and 25 had recently received the influenza vaccine. In total, 150 were enrolled in the standard-of-care arm and 150 in the pay-it-forward arm. All 300 responses were screened for completeness, and were included in the final statistical analyses.

**Fig 1:**
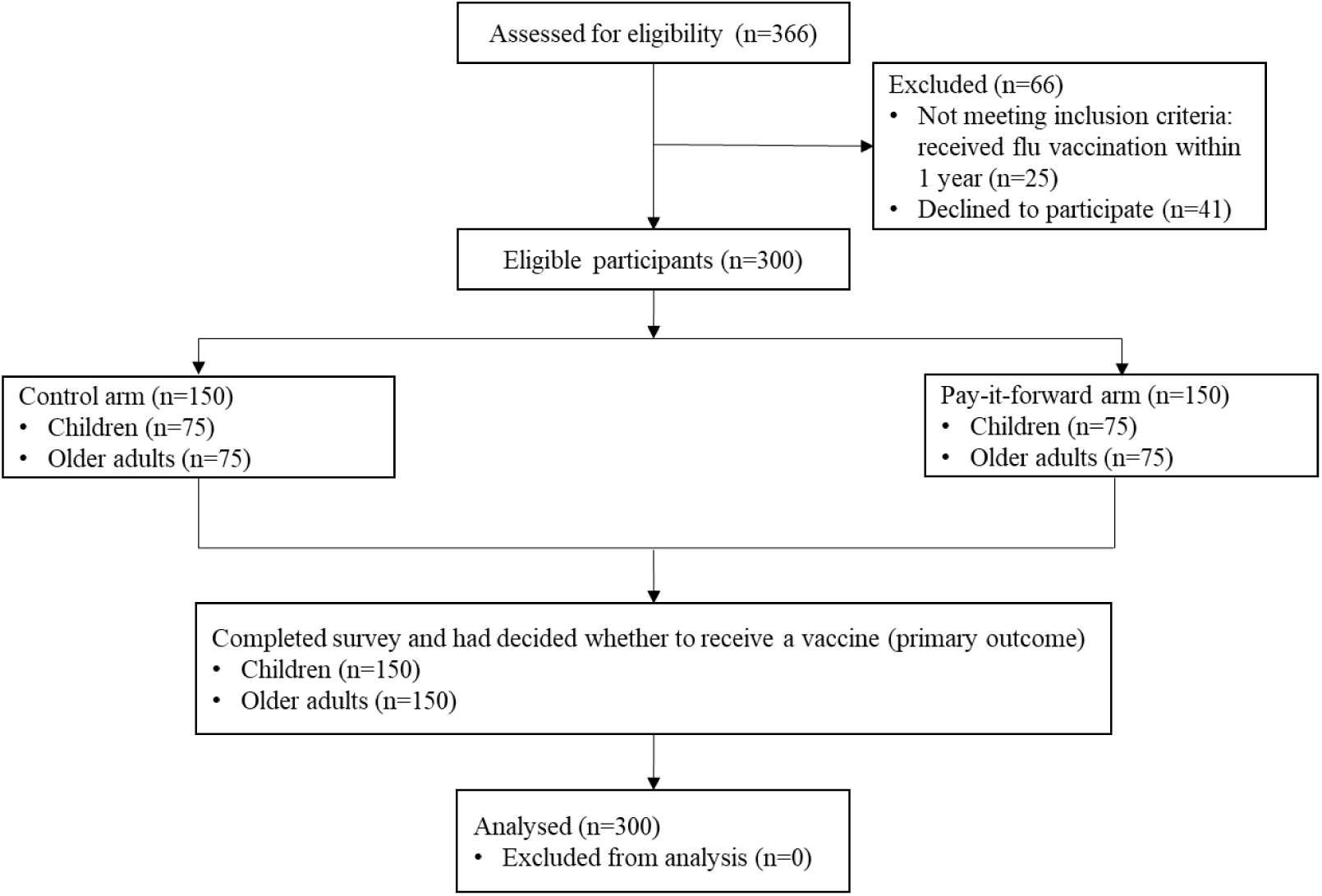
Quasi-experimental study flowchart

Characteristics of caregivers of children and older adult participants were similar between standard-of-care and pay-it-forward arms (Table 1). Regarding the primary outcome of vaccine uptake, 111 (74.0%) of 150 pay-it-forward participants and 55 (36.7%) of 150 participants in the standard-of-care arm received influenza vaccination (χ^2^ test p<0.001; Fig 2; supplementary table 1). Among children, the pay-it-forward arm had an uptake rate of 88.0% (66/75) compared to 53.3% (40/75) in the standard-of-care arm. Among older adults, the pay-it-forward arm had an uptake rate of 60.0% (45/75) compared to 20.0% (15/75) in the standard-of-care arm. Proportion differences in uptake between the pay-it-forward and standard-of-care arms remained statistically significant for both age groups after adjusting for study site and educational level (Supplementary table 1).

**Table 1:** Sample characteristics of recruited child caregivers and older adults in Guangdong Province, China, 2021 (N=300)

|  | Child caregiver group |  | Older adults |  |
| --- | --- | --- | --- | --- |
|  | Standard-of-care<br>(N=75) | Pay-it-forward<br>(N=75) | Standard-of-care<br>(N=75) | Pay-it-forward<br>(N=75) |
| **Age** | 35.91 (10.3) | 36.71 (9.7) | 69.53 (6.4) | 66.52 (6.7) |
| **Sex** |  |  |  |  |
| *Men* | 17 (22.7) | 16 (21.3) | 20 (26.7) | 27 (36.0) |
| *Women* | 58 (77.3) | 59 (78.7) | 55 (73.3) | 48 (64.0) |
| **Education** |  |  |  |  |
| *Elementary school or below* | 8 (10.7) | 4 (5.3) | 33 (44.0) | 26 (34.7) |
| *Middle school* | 45 (60.0) | 26 (34.7) | 31 (41.3) | 42 (56.0) |
| *Undergraduate or above* | 22 (29.3) | 45 (60.0) | 11 (14.7) | 7 (9.3) |
| **Occupation** |  |  |  |  |
| *Unemployed* | 20 (26.7) | 21 (28.0) | 53 (70.7) | 58 (77.3) |
| *Peasant* | 1 (1.3) | 1 (1.3) | 19 (25.3) | 15 (20.0) |
| *Employed* | 54 (72.0) | 53 (70.7) | 3 (4.0) | 2 (2.7) |
| **Annual income (USD)** |  |  |  |  |
| *0-1860* | 19 (25.3) | 23 (30.7) | 38 (50.7) | 28 (37.3) |
| *1860-9300* | 22 (29.3) | 24 (32.0) | 29 (38.7) | 36 (48.0) |
| *9300-1,8600* | 20 (26.7) | 16 (21.3) | 7 (9.3) | 10 (13.3) |
| *>1,8600* | 14 (18.7) | 12 (16.0) | 1 (1.3) | 1 (1.3) |
| **Marital status** |  |  |  |  |
| *Single, divorced, separated or widowed* | 4 (5.3) | 4 (5.3) | 20 (26.7) | 20 (26.7) |
| *Married or living with a partner* | 71 (94.7) | 71 (94.7) | 55 (73.3) | 55 (73.3) |

**Fig 2:**
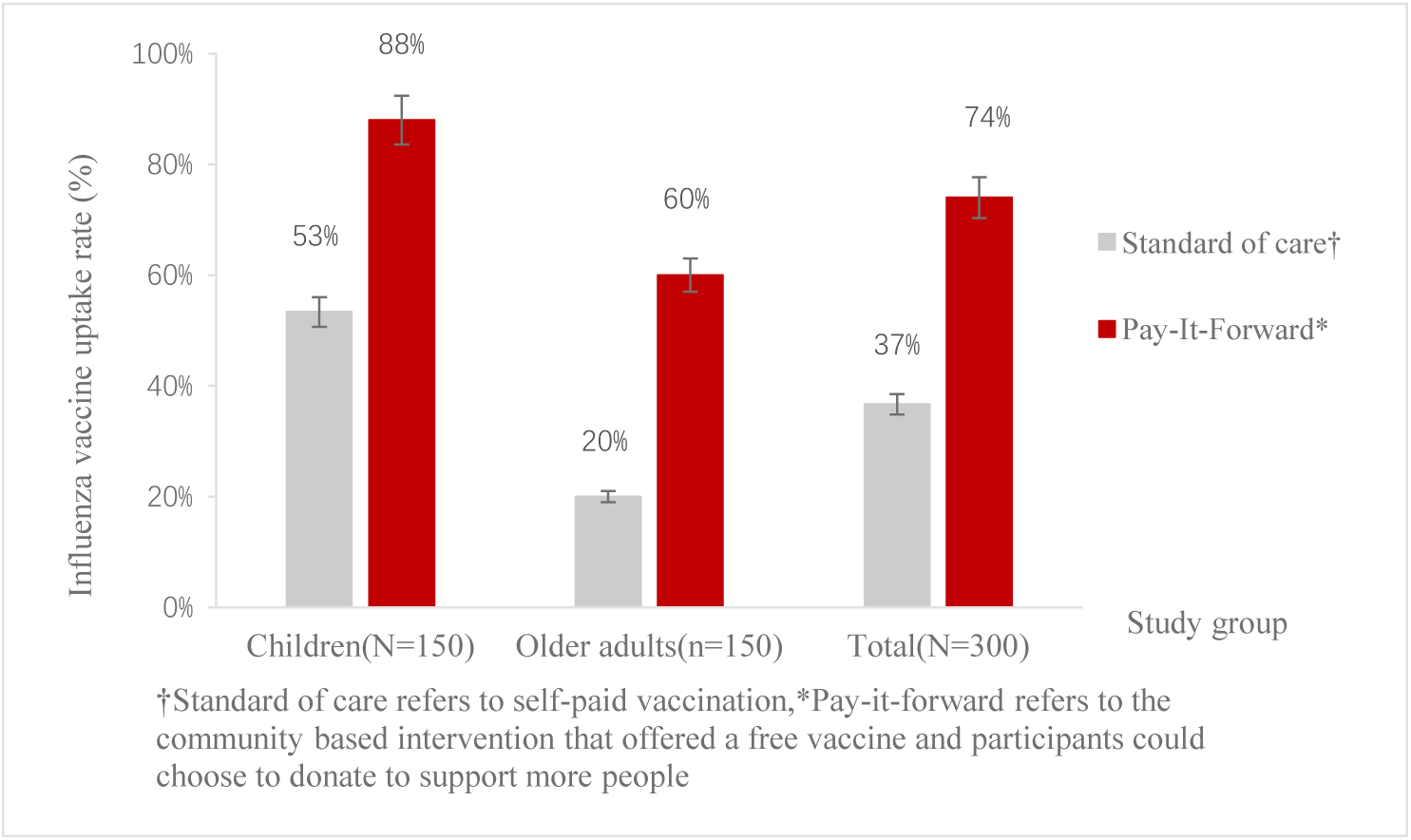
Influenza vaccine uptake rates of standard-of-care and pay-it-forward arms by age group in Guangdong Province, China, 2020-2021 (N=300)

Table 2 suggests that people in the pay-it-forward arm were more likely to receive the vaccine compared to people in the standard-of-care arm (adjusted odds ratio (aOR)= 6.7, 95%CI [2.7, 16.6] among children; aOR=5.0, [2.3, 10.8] among older adults).

**Table 2:**
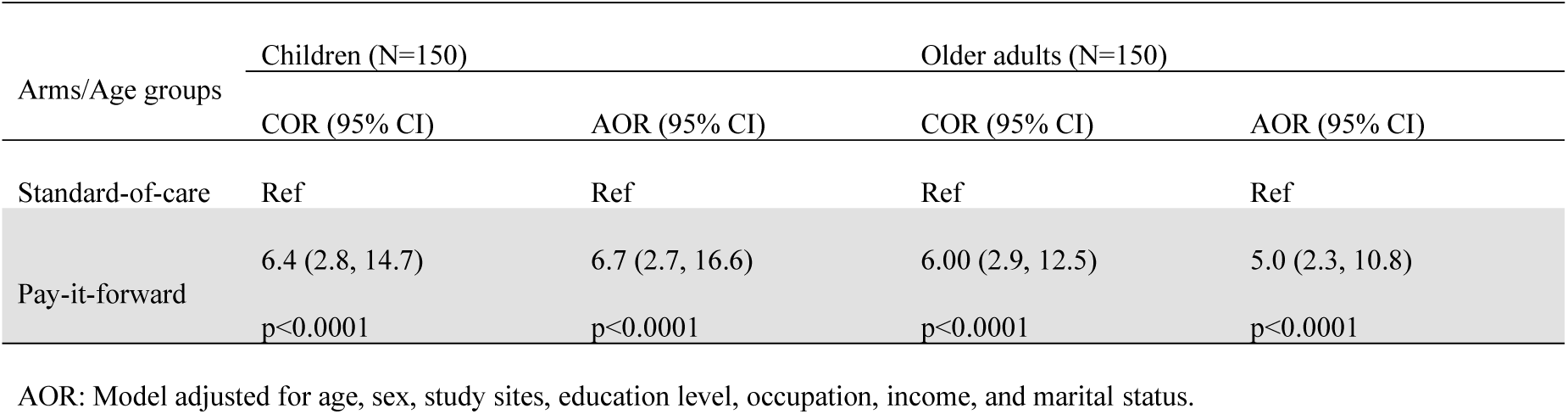
Multivariable logistic regression to compare vaccine uptake rates of standard-of-care and pay-it-forward arms by age groups in Guangdong Province, China, 2020-2021 (N=300)

Confidence levels in safety, importance, and effectiveness were high among the entire sample (75.4%, 78.6%, and 73.7%, respectively). Compared to people in the standard-of-care arm, people in the pay-it-forward arm had higher confidence in influenza vaccine safety (83.3 vs 67.4%; aOR=2.2, [1.2-3.9]), vaccine importance (88.2 vs 68.8%; aOR=3.1, [1.6-6.0]), and vaccine effectiveness (84.7 vs 62.4%; aOR=3.1, [1.7-5.7]) (Table 3).

**Table 3:** Multivariable logistic regression to compare vaccine confidence between standard-of-care and pay-it-forward arms in Guangdong Province, China, 2020-2021 (N=285)

| Study arm | Vaccine confidence - Safety |  |  | Vaccine confidence - Importance |  |  | Vaccine confidence – Effectiveness |  |  |
| --- | --- | --- | --- | --- | --- | --- | --- | --- | --- |
|  | Total | cOR | aOR | Total | cOR | aOR | Total | cOR | aOR |
|  | N(%) | (95% CI) | (95% CI) | N(%) | (95% CI) | (95% CI) | N(%) | (95% CI) | (95% CI) |
| Standard-of-care | 95(67.4) | Reference | Reference | 97(68.8) | Reference | Reference | 88(62.4) | Reference | Reference |
| Pay-it-forward | 120(83.3) | 2.4 (1.4-4.2) | 2.2(1.2-3.9) | 127(88.2) | 3.4 (1.8-6.3) | 3.1(1.6-6.0) | 122(84.7) | 3.3(1.9-5.9) | 3.1(1.7-5.7) |
| *P value* |  | 0.002 | 0.01 |  | <0.001 | <0.001 |  | <0.001 | <0.001 |

284 AOR: Model adjusted for age, sex, occupation, education level, income level

Regarding participant contributions and engagement in the pay-it-forward arm, among 111 participants who received the influenza vaccine, 107 (96.4%) donated money, with a total contribution of US$597.62. Donations covered 36.0% of vaccination costs in the pay-it-forward arm. The median donation was US$4.6. Only 30% of donors in the rural site contributed US$7.6 or above, compared to 61.9% in the suburban and 40.0% in urban sites (supplementary Fig 7 and supplementary table 2). They contributed to the development of influenza vaccination materials, and 19/60 (31.7%) people wrote postcard messages for subsequent participants. Most handwritten messages expressed general good wishes.

The total financial cost of implementing an influenza vaccination intervention for participants was US$2,725 for the standard-of-care arm and US$4,477 for the pay-it-forward arm. The financial cost per person vaccinated was $49.55 for the standard-of-care and $40.33 for the pay-it-forward arm. When economic costs are considered, the economic cost of implementing an influenza vaccination intervention for children and older adults was US$3,557 for standard-of-care and US$5,062 for pay-it-forward. The economic cost per person vaccinated was $64.67 for the standard-of-care and $45.60 for the pay-it-forward arm. The financial and economic costs per person vaccinated in the pay-it-forward arm were close to those ($40.92 and $40.92 respectively) in the free vaccine arm.

## DISCUSSION

Our study contributes to the literature by determining the effectiveness of a social innovation intervention using a quasi-experimental study, developing new methods for public influenza vaccination engagement, and enhancing vaccine uptake. Our data suggest that the pay-it-forward strategy may increase influenza vaccine uptake among high-risk individuals compared to the current self-pay strategy for vaccination. This strategy substantially increased vaccine uptake compared to the standard-of-care, elicited financial contributions, improved vaccine confidence, and co-created participatory messages.

We found that children and older adults who took part in pay-it-forward had higher influenza vaccine uptake than they did if they needed to self-pay for vaccination. This finding is consistent with previous intervention studies using pay-it-forward to improve health services uptake.^13,14^ The vaccination rate in the pay-it-forward arm is also substantially higher than the uptake rate (47.5%) in Chinese cities where influenza vaccination is partially or fully reimbursed.^6^ The effect of pay-it-forward might be related to the reduced costs associated with vaccination, enhanced public engagement and vaccine confidence, or both.

We also observed that, among those enrolled in the pay-it-forward arm, nearly all voluntarily donated to support another person after receiving an influenza vaccine, including those with a low annual income from a study site in a poor rural area. In addition, pay-it-forward had lower costs per person vaccinated than the standard-of-care practice. The financial cost per person vaccinated was also lower than the median cost (US $50.78) per additional enrollee vaccinated from a systematic review published in 2018.^22^ Donations collected using a pay-it-forward system can support more individuals in receiving influenza vaccine services. This may have important implications for effective social innovations targeting improved influenza vaccine uptake while government-funded vaccination is unavailable. Pay-it-forward could potentially transition from out-of-pocket payments to government-funded influenza vaccine programs.

Pay-it-forward has additional social benefits; it generated many messages to drive influenza vaccine uptake. This is a rare example of public engagement in an influenza vaccination program.^10^ Public engagement is central to the success of public health programs; given that some engagement methods could facilitate influenza transmission,^23^ it is especially important to identify public engagement methods that are safe and effective. Engaging the community in vaccination services through cultivating kindness and reciprocity may also strengthen community solidarity and increase confidence in vaccine services.^15,24^

The study has several limitations. First, although our study was implemented after COVID-19 lockdowns were lifted, all sites were heavily focused on COVID-19 prevention, vaccination, and related activities. This caused some delays in recruitment despite the availability of influenza vaccines. But this demonstrates the feasibility of pay-it-forward during emergency responses. Second, we examined people from only three sites. However, our sites all had a high influenza prevalence, included different settings (rural, suburban, urban), and reflected common pathways for vaccination in China. Third, our study did not capture granular data on implementation and was not powered to test differences between the pay-it-forward strategy and the free vaccine arm. More effectiveness research to compare different implementation strategies is needed to differentiate effective components and determine optimal pay-it-forward practices. Finally, this was a quasi-experimental study and did not use randomization. However, all standard of care periods were immediately followed by pay-it-forward periods (supplement 4), decreasing the likelihood of temporal changes explaining the observed differences. In addition, our previous pay-it-forward quasi-experimental study results ^13^ were similar to a subsequent randomized controlled trial. ^14^

Our study has implications for research, implementation, and policy. This study expands the limited literature on public engagement in influenza vaccine programs. It demonstrates how social innovation can engage key communities in the implementation process and build confidence in influenza vaccination. This might help address vaccine hesitancy and anti-vaccine movements. Then, by tapping into community-based primary care clinical settings that provide essential care to the general public, the study also adds to previous pay-it-forward public health research focused on promoting sexually transmitted infectious diseases testing among sexual minorities. This indicates the feasibility of using pay-it-forward in a wider context. The success of the pay-it-forward initiative and different donation levels across three study sites shows the potential to mobilize financial resources between areas with different economic status (e.g., mobilize financial resources from economically better-off areas to subsidize essential preventive services for people in more impoverished areas). Pay-it-forward may also serve as a bridge to public free provision. Future implementation research is needed to better understand the implementation of this system and integrate this intervention within health systems. Pay-it-forward may be particularly relevant in the large number of countries that charge fees for influenza vaccines. Developing pay-it-forward programs could help financially support expanded influenza vaccination programs in these settings while also generating community-engaged messages.

## Data Availability

All data produced in the present study are available upon reasonable request to the authors.

https://drive.google.com/file/d/1vXEVpVSi_x_8P8bva58qL_Wqh7u5bRRB/view?usp=sharing

## ACKNOWLEDGEMENTS

We appreciate support from Dr. David Sarley from Bill & Melinda Gates Foundation for his contribution and support to the early development of the intervention package. We are grateful for comments from Dr. Jing Li at West China School of Public Health Sichuan University to help improve clarity of the paper. We thank Huipeng Liao, Yafei Si, Xu Chen, Xiaolin Qiu, Yuxin Ni, Yuan Xiong, Ruoyu Zhu, Jie Fan, Yumeng Du from University of North Carolina at Chapel Hill Project-China, Fiona Sun and Kaiyi Han from Faculty of Public Health and Policy, London School of Hygiene and Tropical Medicine for their help with questionnaire instrument development, data collection, video production, data visualization and interpretation.

## AUTHOR CONTRIBUTIONS

DW, WT and JDT conceived the idea and designed the study. CJ, ZW, YX, NR, SQ, JB, LW implemented the study and collected data. CJ, KB, FFT, and FJ cleaned and analyzed data, and generated figures, tables to present findings of the paper. JJO, MJ, HJL, TC, LL, WG, and FY provided expert advice on designing the study, analyzing data and interpreting results. SW, WC, YZ, QY and JT coordinated local resources, provided community and stakeholder perspectives to refine the study design, and produced communication materials. DW and CJ wrote the first draft of the paper and all co-authors provided constructive comments and edited the manuscript. All authors have seen and approved the final version of the article.

## DATA AVAILABILITY

Requests for data by researchers with proposed use of the data can be made to the corresponding author with specific data needs, analysis plans and dissemination plans.

## ETHICS APPROVAL

Ethical approval for this study was obtained from the institutional review boards at the London School of Hygiene & Tropical Medicine (approval number 19100) and the Zhuhai Center for Disease Control (approval number 2020011). Online consent was obtained from guardians of children and older adults. The trial was registered in early September 2020 in Chinese Clinical Trial Registry with the number of ChiCTR2000040048.

