## Supplemental files for "Pay-it-forward to improve influenza vaccine uptake and public engagement among children and older adults in China: A quasi-experimental pragmatic trial"

**Supplementary video link**

https://drive.google.com/file/d/1vXEVpVSi_x_8P8bva58qL_Wqh7u5bRRB/view?usp=sharing

**Supplementary Fig 1: Pay-it-forward model overview**

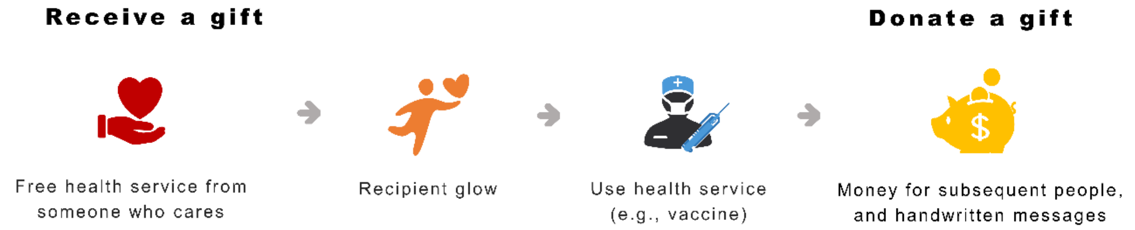


**Supplementary Fig 2 postcard example**

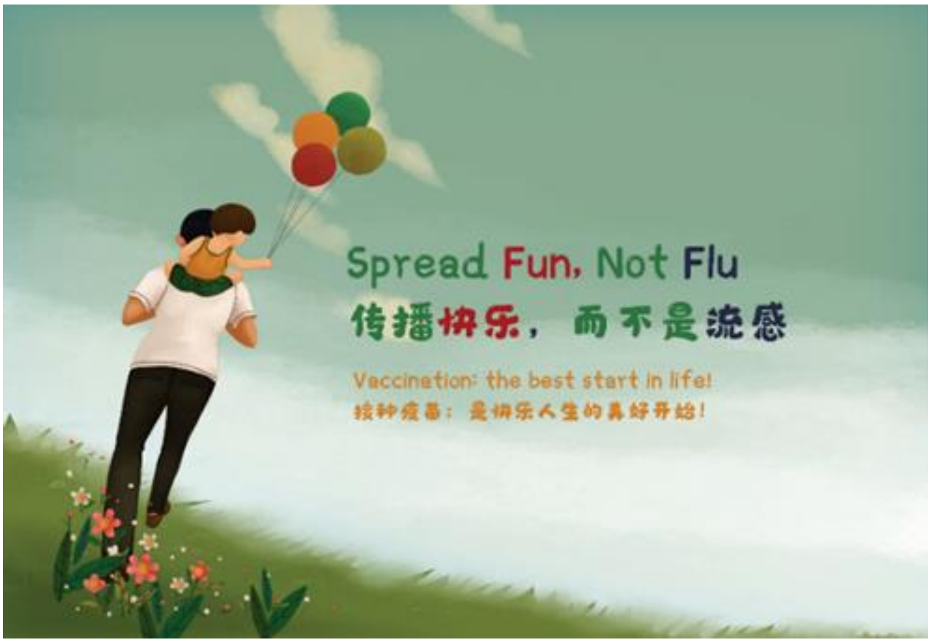

**Supplementary Fig 3: translated hand-written postcard messages**

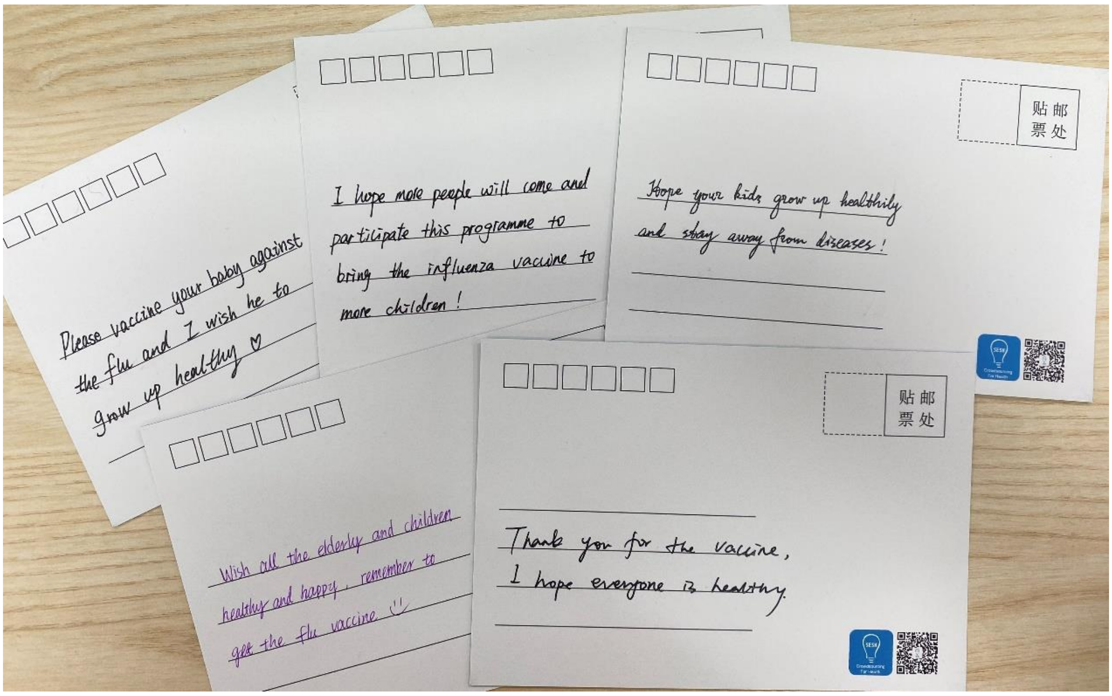


**Supplementary Fig 4 Time-based recruitment for the three study arms at three study sites**

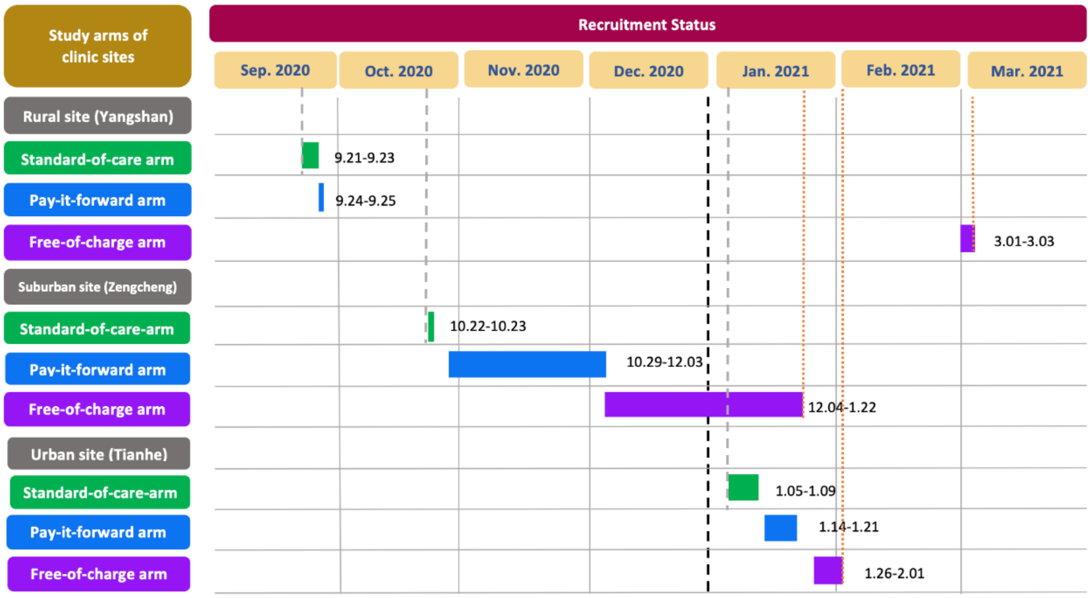


* Each site only used one arm at a single time period and all three sites had standard-of-care arm followed directly by pay-if-forward arm.

Footnote: The recruitment in Yangshan was spaced out because of occasional stock outs and COVID vaccination. The lengths of time for the three arms at the study sites varied due to variations in patient volume and recruitment pace.

### Supplementary Fig 5: Introduction to influenza (English translation).

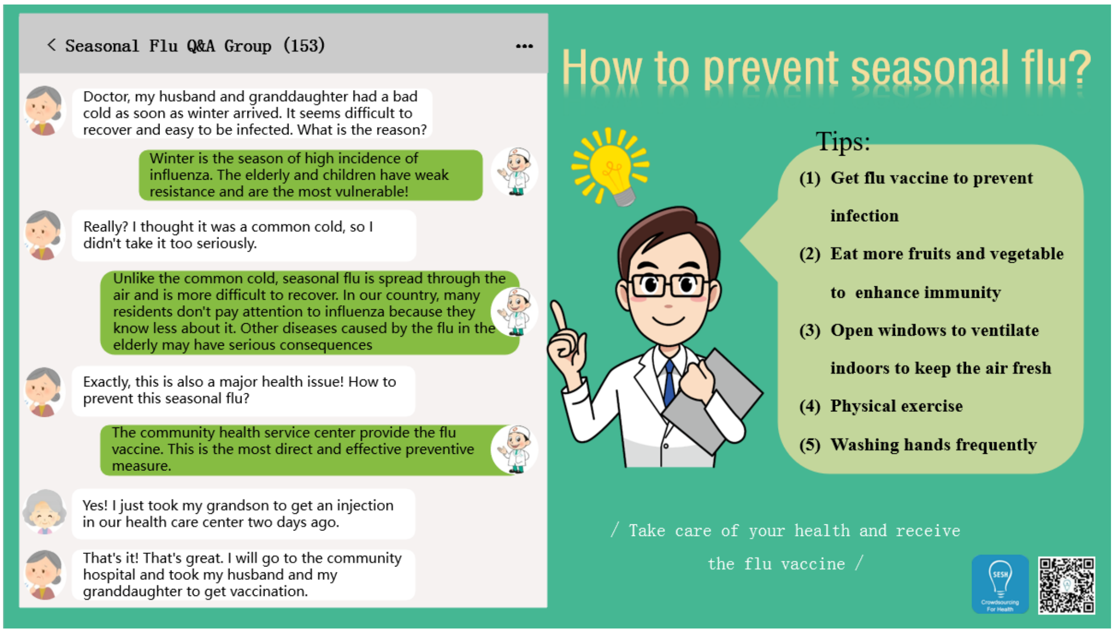


### Supplementary Fig 6: pay-it-forward pamphlet (English translation).

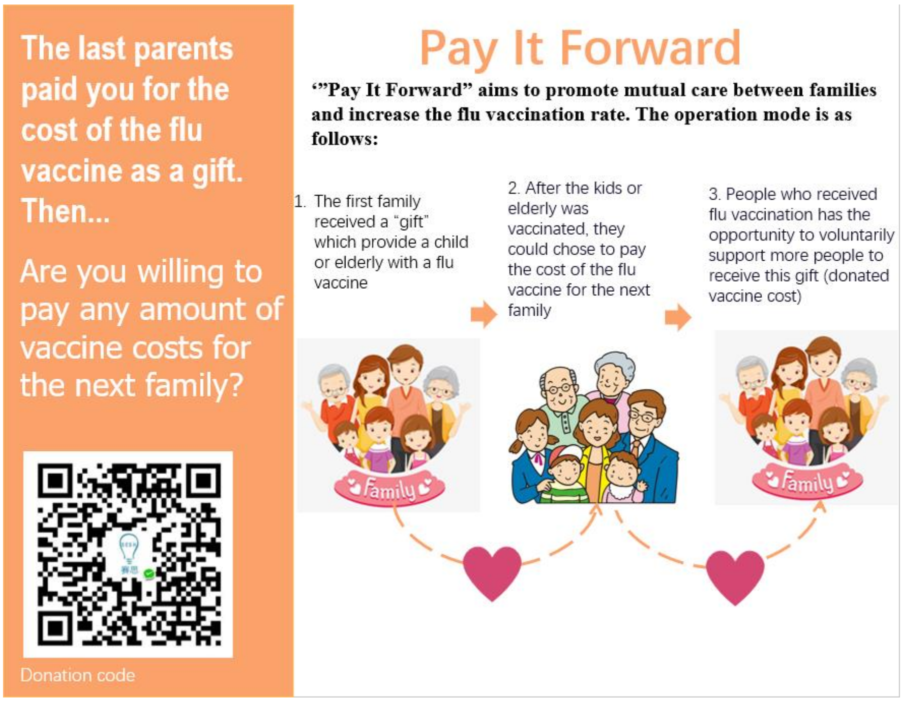

**Supplementary Fig 7: Distribution of donation amounts by study site (n=107)**

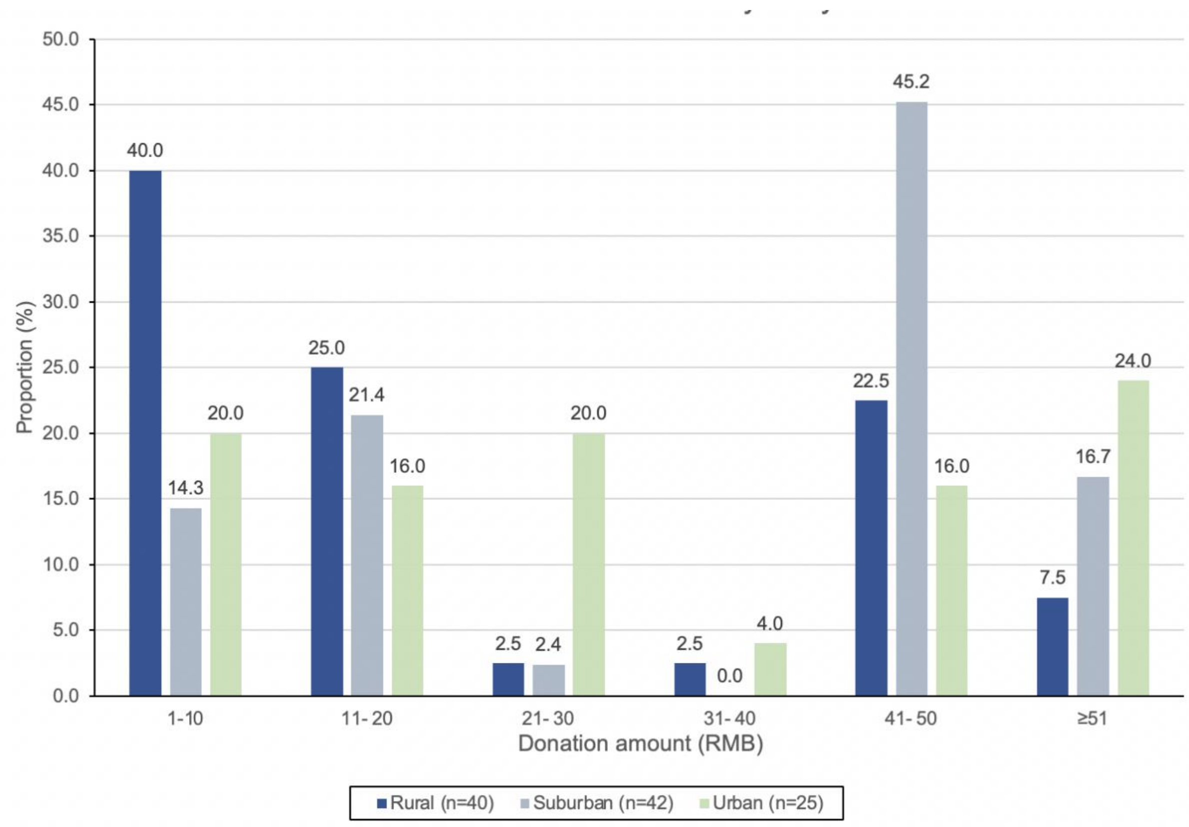


Note: 1 US$=6.59 RMB

Supplementary table 1 Proportion differences in influenza vaccination rates between the three study arms by age groups in Guangdong Province, China, 2020-2021 (N=450)

| | Children<br>N=225 | | Older adults<br>N=225 | |
|---|---|---|---|---|
| **Uptake (n,%)** | | **P value**<br>**<0.0001** | | **P value**<br>**<0.0001** |
| *Standard of care* | 40 (53.3) | | 15 (20) | |
| *Pay-it-forward* | 66 (88) | | 45 (60) | |
| **Study arms** | **Crude proportion difference (95%CI)** | **P value** | **Crude proportion difference (95%CI)** | **P value** |
| *Pay-it-forward vs Standard of care* | 34.7 (21.2, 48.1) | <0.0001 | 40.0 (25.7, 54.3) | <0.0001 |
| **Adjusting study site** | **Adjusted proportion difference (95%CI)** | **P value** | **Adjusted proportion difference (95%CI)** | **P value** |
| *Pay-it-forward vs Standard of care* | 34.7 (21.4, 47.9) | <0.0001 | 40.0 (26.3, 53.7) | <0.0001 |
| **Adjusting education level** | | | | |
| *Pay-it-forward vs Standard of care* | 36.3 (22.1, 50.5) | <0.0001 | 41.1 (27.1, 55.1) | <0.0001 |

Supplementary table 2: Distribution of donation status and participants with a donation of 50 RMB or above by study site, n(%)

| Characteristic | Rural (n=50) | Suburban (n=50) | Urban (n=50) | p-value* |
|---|---|---|---|---|
| **Donation status (n=150)** | | | | |
| - *Donated* | 40 (80.0) | 42 (84.0) | 25 (50.0) | <0.001 |
| - *Did not donate* | 10 (20.0) | 8 (16.0) | 25 (50.0) | |
| **Characteristic** | Rural (n=40) | Suburban (n=42) | Urban (n=25) | p-value* |
| **Donation amount (n=107)** | | | | |
| - *<50 RMB* | 28 (70.0) | 16 (38.1) | 15 (60.0) | 0.013 |
| - *>=50 RMB* | 12 (30.0) | 26 (61.9) | 10 (40.0) | |

Note: US$ 1 = RMB 6.59

**Supplementary costs file**

We compared costs between all three arms. We adopted the same sample size of 150 (75 children and 75 older adults) for the free arm as the other two arms. Participants in the free vaccination arm were invited to participate using the same introductory pamphlet and were provided with free influenza vaccination. They did not receive any community-created messages about the pay-it-forward program. After recruiting all participants for the free arm, we collected costs data (cost file table 1) and analyzed costs (costs file figure 1 and table 2). Economic and financial costs per person are reported in the main text.

**Costs file Table 1:** *Unit costs (in 2020 USD) and frequency of vaccine use*

| Intervention | Cost item | Unit cost | Resource use | Source |
|---|---|---|---|---|
| ***Pay-it-forward*** | | Staff wage per hour (USD/hour)* | | |
| • Start-up costs | | | | |
| | *Time designing postcards to be written on by participants in the PIF programme†* | 7.47 | 1 x 5 hr | Personal communication with research staff |
| | *Time of research fellow participating in preparatory workshop†* | 12.7 | 1 x 5 hr | Personal communication with research staff |
| | *Time of research assistant participating in preparatory workshop†* | 7.47 | 1 x 5 hr | Personal communication with research staff |
| | *Time of nurses participating in preparatory workshop* | 4.81 | 3 x 1 hr | Personal communication with research staff<br>China Social Welfare Foundation (12) |

| | | | | |
|---|---|---|---|---|
| | *Time of clinic coordinators participating in preparatory workshop* | 4.81 | 3 x 1 hr | Personal communication with research staff China Social Welfare Foundation(12) |
| • Fixed costs | | | | |
| | *Vaccinators (doctors) for the three clinics* | 11.07 | 3 x 70 hr | China Social Welfare Foundation (12) |
| • Recurrent costs | | | | |
| | *Time of nurses in recruiting patients to join the PIF programme††* | 4.81 | 3 x 25 hr | Personal communication with research staff<br>China Social Welfare Foundation (12) |
| | *Time of clinic coordinators in performing administrative work for the PIF programme††* | 4.81 | 3 x 25 hr | Personal communication with research staff<br>China Social Welfare Foundation (12) |
| | | Cost per vaccine (USD) | | |
| | *Cost of adult vaccines in Yangshan#* | 22.9 | 41 | Health clinic reimbursement invoices |
| | *Cost of adult vaccine in Zengcheng* | 22.9 | 17 | Health clinic reimbursement invoices |
| | *Cost of child vaccine in Zengcheng* | 8.38 | 25 | Health clinic reimbursement invoices |
| | *Cost of adult vaccine in Tianhe#* | 12.2 | 28 | Health clinic reimbursement invoices |
| | | Cost per item / batch (USD) | | |

| | | | | |
|---|---|---|---|---|
| | *Introductory pamphlets for the PIF programme (batch cost)* | 29.0 | 1 | Project research staff invoices |
| | *Ballpoint pens (for writing messages on PIF postcards) (batch cost)* | 2.25 | 1 | Project research staff invoices |
| | *Cost of printing postcards (batch cost)* | 7.41 | 1 | Project research staff invoices |
| | *Surgical gloves* | 0.075 | 222 | Personal communication with research staff |

| **Intervention** | **Cost item** | **Unit cost** | **Resource use** | **Source** |
|---|---|---|---|---|
| ***Standard-of-care*** | | Staff wage per hour (USD/hour)* | | |
| • Start-up costs | *Time of nurses participating in preparatory workshop* | 4.81 | 3 x 1 hr | China Social Welfare Foundation (12) |
| | *Time of clinic coordinators participating in preparatory workshop* | 4.81 | 3 x 1 hr | China Social Welfare Foundation (12) |
| • Fixed costs | | | | |
| | *Vaccinators for the three clinics* | 11.07 | 3 x 70 hr | China Social Welfare Foundation (12) |
| • Recurrent costs | | | | |
| | *Time of nurses in recruiting patients to receive seasonal influenza vaccination††* | 4.81 | 3 x 12.5 hr | Personal communication with research staff |

| | | | | |
|---|---|---|---|---|
| | | | | China Social Welfare Foundation (12) |
| | *Time of clinic coordinators in performing administrative work for influenza vaccination††* | 4.81 | 3 x 12.5 hr | Personal communication with research staff<br>China Social Welfare Foundation (12) |
| | | Cost per vaccine (USD) | | |
| | *Cost of adult vaccines in Yangshan#* | 22.9 | 15 | Health clinic reimbursement invoices |
| | *Cost of adult vaccine in Zengcheng* | 22.9 | 5 | Health clinic reimbursement invoices |
| | *Cost of child vaccine in Zengcheng* | 8.38 | 14 | Health clinic reimbursement invoices |
| | *Cost of adult vaccine in Tianhe#* | 12.2 | 21 | Health clinic reimbursement invoices |
| | | Cost per item (USD) | | |
| | *Surgical gloves* | 0.075 | 110 | Personal communication with research staff |

| Intervention | Cost item | Unit cost | Resource use | Source |
|---|---|---|---|---|
| ***Free vaccination*** | | | | |
| • Start-up costs | | Staff wage per hour (USD/hour)* | | |
| | *Time of nurses participating in preparatory workshop* | 4.81 | 3 x 1 hr | Personal communication with research staff<br>China Social Welfare Foundation (12) |
| | *Time of clinic coordinators participating in preparatory workshop* | 4.81 | 3 x 1 hr | Personal communication with research staff<br>China Social Welfare Foundation (12) |

| | | | |
|---|---|---|---|
| • Fixed costs | | | |
| *Vaccinators for the three clinics* | 11.07 | 3 x 70 hr | China Social Welfare Foundation (12) |
| • Recurrent costs | | | |
| *Time of nurses in recruiting patients to join the PIF programme††* | 4.81 | 3 x 12.5 hr | Personal communication with research staff<br>China Social Welfare Foundation (12) |
| *Time of clinic coordinators in performing administrative work for the PIF programme††* | 4.81 | 3 x 12.5 hr | Personal communication with research staff<br>China Social Welfare Foundation (12) |
| | Cost per vaccine (USD) | | |
| *Cost of adult vaccines in Yangshan#* | 22.9 | 42 | Health clinic reimbursement invoices |
| *Cost of adult vaccine in Zengcheng* | 22.9 | 13 | Health clinic reimbursement invoices |
| *Cost of child vaccine in Zengcheng* | 8.38 | 18 | Health clinic reimbursement invoices |
| *Cost of adult vaccine in Tianhe#* | 12.2 | 41 | Health clinic reimbursement invoices |
| | Cost per item / batch (USD) | | |
| *Surgical gloves* | 0.075 | 228 | Personal communication with research staff |
| *Information pamphlet for free vaccination arm* | 20.4 | 1 | Project research staff invoices |

***PIF*** *- pay-it-forward,* ***USD*** *– United States dollars*

**Hourly wages were calculated from monthly or yearly wages based on an assumption of 250 working days per year and 10 working hours per day.*

*†These costs were annualised over a three-year period at a discount rate of 3% as they were part of the preparatory workshop for the conception of PIF programme specifically. The training, supplies and expertise gained through these sessions are expected to be useful in future years to inform further iterations of the programme and were therefore annualised over three years.*

*†† It was assumed that each nurse spent 10 minutes per patient to recruit and persuade participants to be vaccinated in the pay-it-forward programme, while they spent 5 minutes per patient for the standard-of-care group, and 5 minutes per patient in the free vaccination arm.*
*#Only adult influenza vaccine was used in Yangshan and Tianhe, i.e. children were inoculated with the same influenza vaccine as the adults in those study sites.*

*Addendum: Breakdown for obtaining the total economic cost and the total financial cost of each intervention arm*

**Total economic cost** of an intervention
= Start-up costs + Fixed costs + Recurrent costs

**Total financial cost** of an intervention
= Start-up costs + Fixed costs + Recurrent costs – (Payment / Donation contributions)

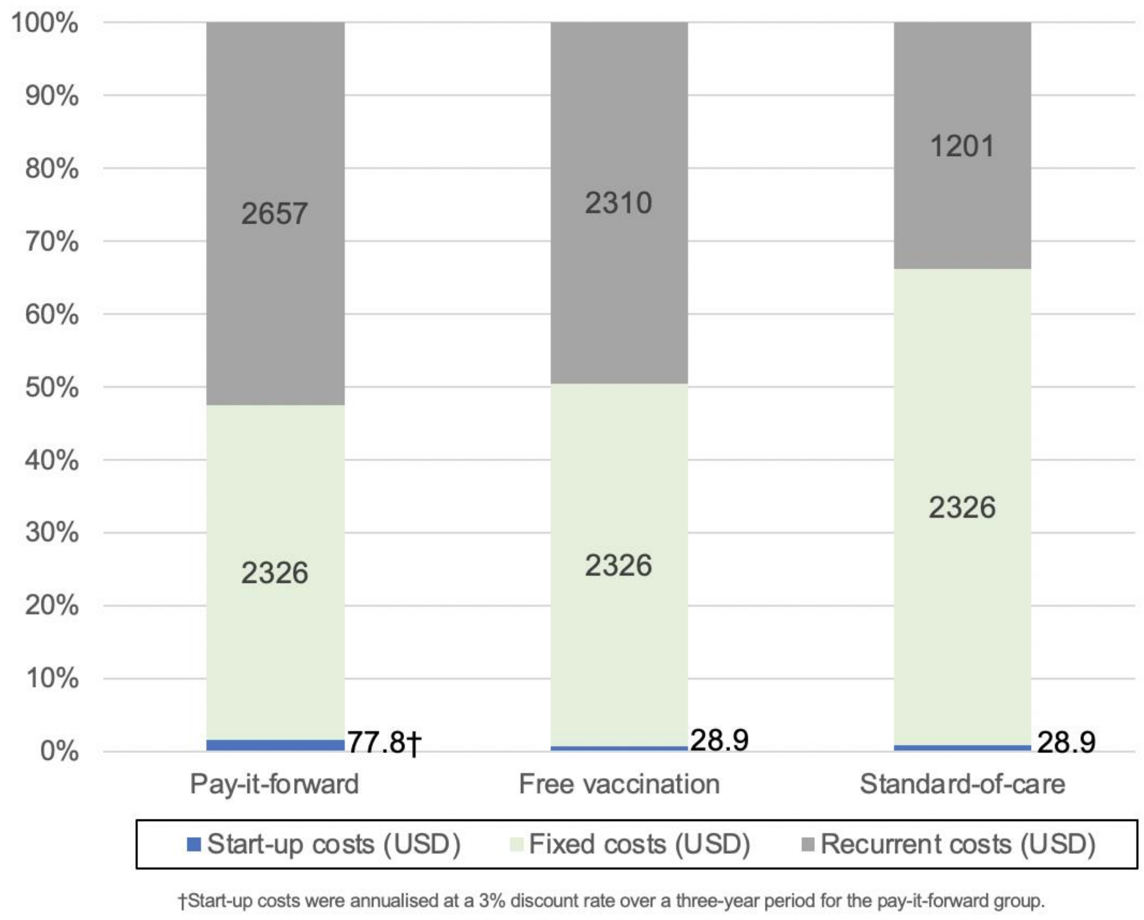


*Costs file Figure 1: Breakdown of economic costs by category in the three intervention arms*

**Costs file** *Table 2: Total economic costs and total financial costs by intervention arm*

| Intervention arm | Total economic cost (USD) | Total financial cost (USD) |
|---|---|---|
| Pay-it-forward | 5062 | 4477 |
| Free vaccination | 4665 | 4665 |
| Standard-of-care | 3557 | 2725 |

*Note: See Table 1 for detailed breakdown of the economic and financial costs*

**Supplementary questionnaire**

**Influenza vaccination for children and elderly people**

**About this study:**
You are invited to participate in this influenza vaccination study. The results of this study can provide evidence for the government to develop better intervention strategies to promote influenza vaccination in China and prevent seasonal influenza among at-risk populations.

**Details of this study:**

You will need to fill out a questionnaire. This questionnaire will ask about your socio-demographic information and opinions towards influenza vaccine. To protect your privacy, all your answers will be transformed into digital formats and password encrypted. If you are eligible to participate in this program, out project personnel will provide you with more detailed information. To confirm whether you are qualified to participate in this program, we will first ask you a few screening questions. If you are not eligible for this program, we will not keep your survey information.

**Note:** If the participant is a child, the guardian will need to consent on the child's behalf and fill in the questionnaire. For the purpose of equality, only one quota is provided for each participating family, either for a child or an elderly individual.

**Screening questions (all groups)**

i.Which category do you belong to____

○Caregivers of a 3-8 year-old children

○Elderly people (>=60)

ii. Your age (fill in a number) ____

iii. Your gender（male-1, female-2）___

iv. Have you been vaccinated against influenza in the past year?

○Yes (Please skip to the end of the questionnaire and submit the answer sheet)

○No

**A. Basic information (all groups)**

1. Ethnicity

○Han

○Other, please specify： ____________

2. Your highest education level:

○Primary school
○Junior middle school

○High school

○Undergraduate or college

○Postgraduate or above

3. Your occupation is:

○Student

○Civil servant

○Farmer

○Ordinary workers (blue collar)

○Company staff (white collar)

○Technical personnel

○Unemployed or retired

○Other： ___________________*

4. Your personal monthly income:

○0-1000 RMB/Month

○1000-5000 RMB/Month

○5000-10,000 RMB/Month

○10,000 RMB/Month or above

5. Your current legal marriage status is:

○Unmarried

○Engaged or married

○Separated or divorced

○Widowed (skip questions 6-9)

6. Age of your partner： ____

7. The highest education level of your partner:

○Primary school

○Junior middle school

○High school

○Undergraduate or college

○Postgraduate

8. The occupation of your partner is:

○Student

○Civil servant

○Farmer

○Ordinary workers (blue collar)

○Company staff (white collar)

○Technical personnel

○Unemployed or retired

○Other： ___________________*

9. Your partner's personal monthly income

○0-1000 RMB/Month

○1000-5000 RMB/Month

○5000-10,000 RMB/Month

○10,000 RMB/Month or above

10 Do you have children in your family?

○Yes

○No (skip question16)

11. How many children are there in your family? (fill in your children's age and sex in chronological order, for example, Child 1, 2 years old, male-1 or

female-2)

○Child 1

Age_____

Sex_____

○Child 2

Age_____

Sex_____

○Child 3

Age_____

Sex______

○Child 2

Age_____

Sex_____

○Child 2

Age_____

Sex_____

12. Do you have elderly people in your family?

○Yes

○No

13. How many elderly people (>=60 years) are there in your family? Age and gender?

○Elderly individual 1

Age:

Sex:

○Elderly individual 2

Age:

Sex:

**B-0. “Standard of care” group**

1. Main purpose of coming to the clinic？

○See a doctor for myself

○Take my child(ren) to see a doctor

○Take my child(ren) to vaccinate

○Accompany elderly family members to see a doctor

○ Body examination

○Other： ____________________ *

2. The researcher has introduced the influenza vaccine. Would you like to participate in the program to vaccinate your child (or yourself if older than 60 years) today?

○Yes

○No（Please skip to the question 22）

3. Who would you like to vaccinate today?

○My Child

○Elderly family member

4.The information of the vaccination recipient?

○Sex___

○Age___

5. Has anyone in your family got seasonal flu in the past year?

○Yes

○No（Skip to the Question 24）

6.If yes, who got seasonal flu in your family?

○Children

○Elderly people

○Children’s parents

7. If you want your family to vaccinate, what is the main reason?

○Children in my family are easy to get seasonable flu

○Elderly people in my family are easy to get seasonable flu

○Both my children and the old people in my family are easy to get seasonable flu

○I’m easy to get seasonable flu

○Recommended by my friends/family member

○Recommendation by HCP the clinic

○Other, please note： ______________ *

8. If you don't want to vaccinate your child or an elderly individual in your family today, why not? [multiple choices] * (if you answered "yes" to question 2, skip this item)

□I don't know enough about seasonal influenza

□I'm not sure about the effect of the influenza vaccine

□My family don't need vaccinations

□It’s too cumbersome

□I'm worried about the side effects

□ I think it is too expensive

□Other, please specify: _______________*

**B-1. “Pay it forward” group**

1. Main purpose of coming to the clinic？

○See a doctor for myself

○Take my child(ren) to see a doctor

○Take my child(ren) to vaccinate

○Accompany elderly family members to see a doctor

○Other： ____________________ *

2. The researcher has introduced the influenza vaccine and ‘Pay it forward’ program to you. Would you like to participate in the program to vaccinate your child (or yourself if older than 60 years) today?

○Yes

○No（Please skip to the question 27）

3. Who would you like to vaccinate today?

○My Child

○Elderly family member

4.The information of the vaccination recipient?

○Sex___

○Age___

5. Are you willing to donate some money to the next family to get the same influenza vaccination?

○Yes

○No（Skip to the Question 25）

6. How much would you like to donate to the next family?

○200 RMB (support one family to get vaccination (1elderly+1children))

○150 RMB (support 1 elderly or 3 children get vaccination)

○100 RMB (support 2 children get vaccination)

○50 RMB (support 1 children to get vaccination)

○Other amount_____

7. If you want your family to vaccinate, what is the main reason?

○Children in my family are easy to get seasonable flu

○Elderly people in my family are easy to get seasonable flu

○Both my children and the old people in my family are easy to get seasonable flu

○I'm easy to get seasonable flu

○Recommended by my friends/family member

○Recommendation by medical personnel at the clinic (irrelevant to 'Pay it forward' program)

○The 'Pay it forward' program

○Other, please note：______________ *

8. What do you think are the benefits of the 'Pay it forward' program?

[Multiple choice]

□Other families' donation can lower my financial burden

□I learn about influenza vaccines that can prevent flus

□Can promote more families to be vaccinated against influenza

□Can reduce the spread of influenza

□Spread love and warmth within the community

□Other：_______________________ *

9. If you don't want to vaccinate your child or an elderly individual in your family today, why not? [multiple choices] * (if you answered "yes" to question 2, skip this item)

□I don't know enough about seasonal influenza

□I'm not sure about the effect of the influenza vaccine

□My family don't need vaccinations

□It’s too cumbersome

□I'm worried about the side effects

□Other, please specify: _______________*

**B-2. “Free vaccination” group**

1. Main purpose of coming to the clinic？

○See a doctor for myself

○Take my child(ren) to see a doctor

○Take my child(ren) to vaccinate

○Accompany elderly family members to see a doctor

○ Body examination

○ elderly get vaccination

○Other：____________________ *

2. The researcher has introduced the influenza vaccine and free vaccination programme. Would you like to participate in the program to vaccinate your child (or yourself if older than 60 years) today?

○Yes

○No（Please skip to the question 27）

3. Who would you like to vaccinate today?

○My Child

○Elderly family member

4.The information of the vaccination recipient?

○Sex___

○Age___

5. Has anyone in your family got seasonal flu in the past year?

○Yes

○No（Skip to the Question 25）

6.If yes, who got seasonal flu in your family?

○Children

○Elderly people

○Children’s parents

7. If you want your family to vaccinate, what is the main reason?

○Children in my family are easy to get seasonable flu

○Elderly people in my family are easy to get seasonable flu

○Both my children and the old people in my family are easy to get seasonable flu

○I’m easy to get seasonable flu

○Recommended by my friends/family member

○Recommendation by HCP at the clinic

○Because the free vaccination programme

○Other, please note： ______________ *

8. If you don't want to vaccinate your child or an elderly individual in your family today, why not? [multiple choices] * (if you answered "yes" to question 2, skip this item)

□I don't know enough about seasonal influenza

□I'm not sure about the effect of the influenza vaccine

□My family don't need vaccinations

□It’s too cumbersome

□I'm worried about the side effects

□Other, please specify: _______________*

**C. Vaccine related information (all groups)**

1. Have you heard about the flu vaccine before this program?

○Yes ○No

2. In general, I think influenza vaccine is important. [Single choice] *

○Strong disagree ○Disagree ○Agree ○Strongly agree

3. In general, I think the influenza vaccine is safe. [Single choice] *

○Strong disagree ○Disagree ○Agree ○Strongly agree

4. In general, I think the influenza vaccine is effective. [Single choice] *

○Strong disagree ○Disagree ○Agree ○Strongly agree

5. Has your child ever been vaccinated against influenza?

(if there is no child in your family, please skip this question) [Single choice] *

○Yes ○No

6. Have elderly people in your family ever been vaccinated against influenza

(if there are no elderly individuals in your family, please skip this question) [Single choice] *

○Yes ○No

7. Have you ever been hesitant about having your child or elderly family members to get the flu vaccine (except for allergies)? [Single choice] *

○Yes ○No

8. Have you ever postponed your child or elderly family members to get the flu vaccine (except for allergies)? [Single choice] *

○Yes ○No

9. Have you ever been refused to have your child or elderly family members to get the flu vaccine (except for allergies)? [Single choice] *

○Yes ○No

10. Have you ever heard about negative information about influenza vaccine?

○Yes ○No

11. Do any of your friends or relatives object to the influenza vaccination?

○Yes ○No

12. Have you or people around you had adverse reactions to influenza vaccination?

○Yes ○No

13. Do you trust the advice provided by the medical personnel in the clinic on influenza vaccine?

○Yes ○No

14. Is price of the vaccine a barrier for your child and/or elderly individuals in your family to get the influenza vaccine?

○Yes ○No

**Supplementary file: protocol versions**

### Summary of Protocol Changes

1. Version 1, Jan 28, 2020. This was the pilot version used for conducting a feasibility pilot as well as for sponsorship application from LSHTM (please see the sponsorship letter at the end of this file). The initial proposed sample size of 600 was a rough estimate based on previous observational studies and for three interventions arms in two vaccination clinics (one in a developed areaand one in an underdeveloped area) in Guangdong province.
2. Version 2, April 14, 2020. This was the version used for a revised LSHTM IRB application and approved by LSHTM. We started to establish connectionswith local community-based health organizations and vaccination sites, and tentatively decided on three study sites which included one in Guangzhou city,one in Yangshan county and one in Shenzhen city. Given Shenzhen is a city that is providing free vaccines to the two target age groups, we tentatively decided to recruit all participants for free vaccine treatment in Shenzhen as an exploratory arm. The flowchart was therefore updated accordingly. We also conducted a feasibility pilot test and then calculated a sample size based on the pilot results. We adjusted the total sample size from 600 to 300, with 100 for each arm. We added a section on sample size calculation and clarified that the sample size is calculated to primarily test differences in uptake between pay-it-forward and standard of care arms but not between pay-it-forward and free arms.
3. Version 3, February 9, 2021. We expanded the sample size to 450 (50 more participants for each arm) to allow sub-analysis by study site and age group. Shenzhen study site was removed because of logistic difficulties. Instead, we confirmed availability of influenza vaccine at an alternative suburban site in Zengcheng City and revised the recruitment plan of participants for free vaccine treatment at all three study sites. We added a section on intervention development.

We revised the order of recruitment for the three treatment groups as standard of care, pay-it-forward, and free vaccination. We clarified the secondary outcomes (factors associated with vaccine uptake, vaccine confidence and hesitancy, and economic evaluation), revised our hypotheses and provided more information on the data analysis plan. We added the China clinical trial number.

Research Proposal Version No.: 1

Research Proposal Version Date: Jan 28, 2020

**Title: Pay-it-forward to improve influenza vaccine uptake among children andolder people in China: A quasi-experimental study**

**Trial Sponsor**

Name: London School of Hygiene & Tropical Medicine

Address: London School of Hygiene & Tropical Medicine, Keppel Street, London WC1E7HT

**Project Summary**

Despite a high burden of influenza-related mortality, China has extremely low rates of influenza vaccination among key groups including children and older people. Ten people die of influenza each hour in China. Individuals ≥60 years is 26 times more like to die of influenza than people < 60. Kids (≤5 years) also have extremely high influenza related death. However, only 2% of Chinese are vaccinated. Influenza vaccines cost 9-16 GBP and are not covered by national insurance programs. We propose a pay-it-forward intervention to increase influenza vaccination rates among children aged between 6months and 8 years, as well as older people aged 60 or above in community-based health organizations. Pay-it-forward has one person receive a gift (e.g., a free influenza vaccination to a caregiver of a child or an old individual) and then asks them if theywould like to pay-it-forward for other individuals to benefit from the same generous gift. Participants will also be able to create handwritten postcards encouraging people in the local community to pay-it-forward to the next individual and donate to the rolling finance pool. We will evaluate the intervention using a quasi-experimental study including three conditions (ie, pay-it-forward, free of charge, and pay out-of-pocket) at two community health organizations in Guangdong province, Southern China – one from a developed city and the other one from an under-developed county. We will recruit 100 children and 100 older individuals (50 children and 50 older individuals from each community health organization) respectively during the pay-it-forward condition period. Likewise, the expected sample sizes for the other two conditions will be the same. A total of 600 participants will be recruited, including 300 children (consented by the guardian) and 300older individuals. Primary outcomes are influenza vaccine uptake rates and pay-it-forward donation rate.

**Background**

Influenza is a viral respiratory infectious disease of global importance. Seasonal influenza epidemics cause 3 to 5 million severe cases and 290,000 to 650,000 deaths in the world each year.[1] Pregnant women, infants and children, older people, and patients with chronic diseases are at high risks of serious illness and death when infected. In China, about 10 people die from influenza-related illnesses each hour. [2] People older than 60 years old are26 times more like to die from influenza than younger individuals.[2] Influenza vaccinationis considered the most effective way to prevent influenza-related diseases. For example, globally, over 40% of countries have decided to provide free flu vaccine to key populations such as children and older individuals.

Chinese Center for Diseases Control and Prevention (China CDC) has released in 2018 a high-level guideline for influenza vaccination services provision to at-risk populations and provinces are developing piloting programs. The guideline defines several at-risk populations including pregnant women, children, family members and caregivers ofinfants less than 6 months of age, and people aged over 60 years old. However, the most recent pooled evidence revealed that only 11.9% of children (aged 6 months to 17 years old) and 21.7% of older population (aged >=60 years) were vaccinated in China.[3]

There are several reasons for low influenza vaccination rates in the country. First, influenza vaccines are not subsidized by government funding in most places due to governmental financial constraints and people need to pay around 16 pounds out-of-pocket for getting the vaccination. Second, the lack of trust in vaccines generally likely decreases demand for influenza vaccine in

China. In July 2018, a massive vaccine scandal swept across China, and over 500,000 children were likely injected with faulty triple vaccines (diphtheria/pertussis/tetanus, DPT).[4] This scandal dramatically decreased trust towards vaccine programs, including influenza vaccine. In addition, pregnancy is widely considered a contraindication for influenza vaccination, and pregnant women in many places are rejected by health workers for vaccination.[5] In places where influenza vaccines are free, public awareness and vaccination rates are suboptimal compared to other countries.[6] The vast majority of the population are unaware of the vaccination and there is widespread hesitancy in safety and effectiveness of influenza vaccine, even among health workers.[7] Only 8% of health workers always recommended vaccine to patients during the influenza season.[7] Innovative strategies are needed to reduce financial burden, build up public trust and collect evidence on effective interventions to improve vaccine uptake.

Pay-it-forward is an innovative, participatory behavioral intervention. Pay-it-forward has one individual receive a free influenza vaccine who will be informed by using hand-written postcard messages co-created by participants that someone else paid for them, then ask if they would like to support a future vaccination. Figure 1 illustrates how the model works. Our previous pay-it-forward studies focused on diagnostic services uptake among marginalized populations have proven effective in increasing chlamydiaand gonorrhea tests uptake by three folds, compared to the control group.[8] In addition,89%of participants donated to the rolling finance pool and the community generosity build up trust in health services.[8] Evidence also showed simple acts of kindness are contagious [9] and hold potential to be leveraged to enhance public health services delivery.

In this quasi-experimental study, we propose a pay-it-forward approach, aiming to improve public trust and influenza vaccine uptake among two key sub-groups, namely children aged between 6 months and 8 years old and older people aged 60 or above in Guangdong province. We determined these two sub-groups based on the disease burden and the national definition of at-risk populations. Specific objectives include 1) to examine the feasibility of applying the pay-it-forward approach in influenza vaccine research in the community setting; 2) to compare the effectiveness of three strategies, which are pay-it-forward, free of charge, and pay out-of-pocket (standard care condition), on influenza vaccine uptake among children and older individuals in China; 3) to investigate the donation rate and average donation amount among participants in the pay-it-forward condition.

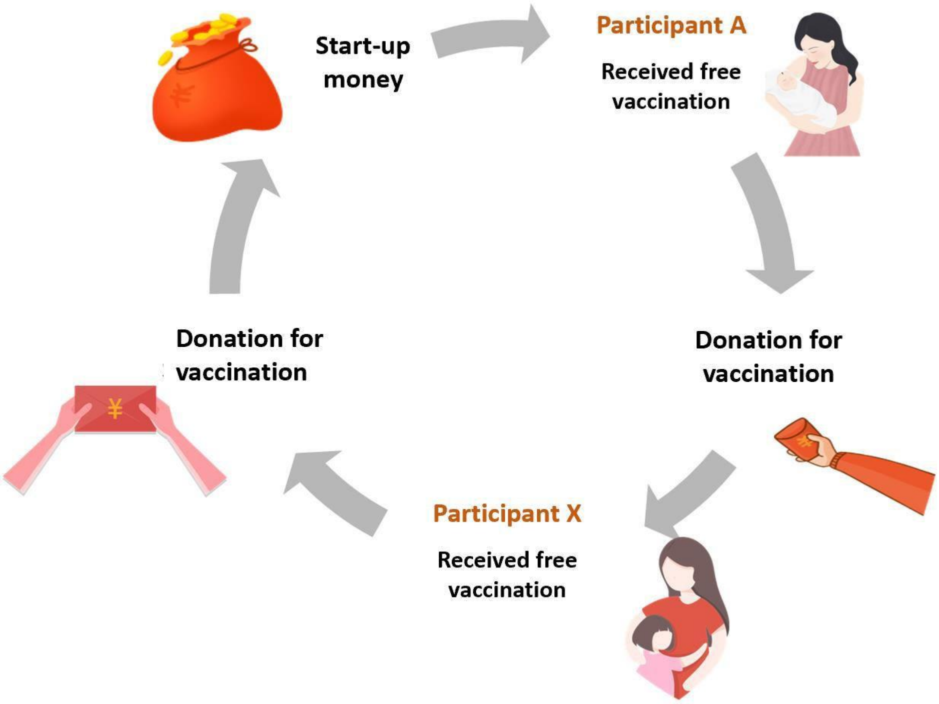


Figure 1: "Pay-it-forward" Model

**Hypotheses**

Based on the previous research, we conservatively estimate that compared to the standardcare (out-of-pocket pay) group,

1) "pay-it-forward" model can significantly improve the influenza vaccination rate among at-risk population including children and older groups (30% higher than the standard care group)
2) The donation rate of participants in the "pay it forward" group can reach 50% or higher
3) The "pay it forward" donation can cover 10% of the total economic cost of influenza vaccine.

**Methods**

*Study setting*

Guangdong is a subtropical southern province with a population over 100 million in China. In southern China, influenza is prevalent throughout the year; it has a clear peak inthe summer and a less pronounced peak in the winter, including Guangdong.[10] In response to the national guideline to provide influenza vaccine to at-risk populations, Guangdong Province has released relevant provincial level policies to promote influenza vaccine. Stockouts of influenza vaccines have been reported in different regions due to tighter quality control of pharmaceutical products after the national vaccine scandal in 2018.[11] Therefore, identifying study sites with sufficient vaccine supply is key to the success of the pilot study during the post-scandal transitioning period. Two batches of tetravalent influenza vaccines produced by Hualan Biology company have been issued in August, 2019; and the first 500,000 tetravalent influenza vaccines were supplied to eight provinces in China, including Guangdong. Given above factors, Guangdong province is selected as the major study location. The London School of Hygiene and Tropical Medicine (LSHTM) research team has established a local collaborative institute with

strong primary care networks in the province - Guangdong Second Provincial General Hospital.

*Target population and inclusion criteria*

The pilot study will focus on children and older people. The inclusion criteria of this study are divided into two age groups:

1） Children: i) children aged between six-month-old and eight years old, ii) no acute moderate or severe illnesses with or without fever, iii) no severe, life-threatening allergies to flu vaccine or any ingredient(s) in the vaccine., iv) a legal guardian, be it a parent or grandparent in the Chinese setting, consents to participate the study.

2） Older people: i) ≥ sixty years old, ii) no acute moderate or severe illnesses with or without fever, iii) no severe, life-threatening allergies to flu vaccine or any ingredient(s) in the vaccine, iv) intelligently capable of making informed decisions and consent to participate the study.

*Study design*

In this study, two vaccination clinics (one in developed and one in underdeveloped areas) will be selected in Guangdong Province for recruitment. The selected community health centers or vaccination clinics should have established infrastructure to provide influenza vaccination services, e.g. have sufficient influenza vaccine stock, relevant medical personnel including designated physicians and nurses for providing vaccine services. Theresearch design is shown in Figure 2.

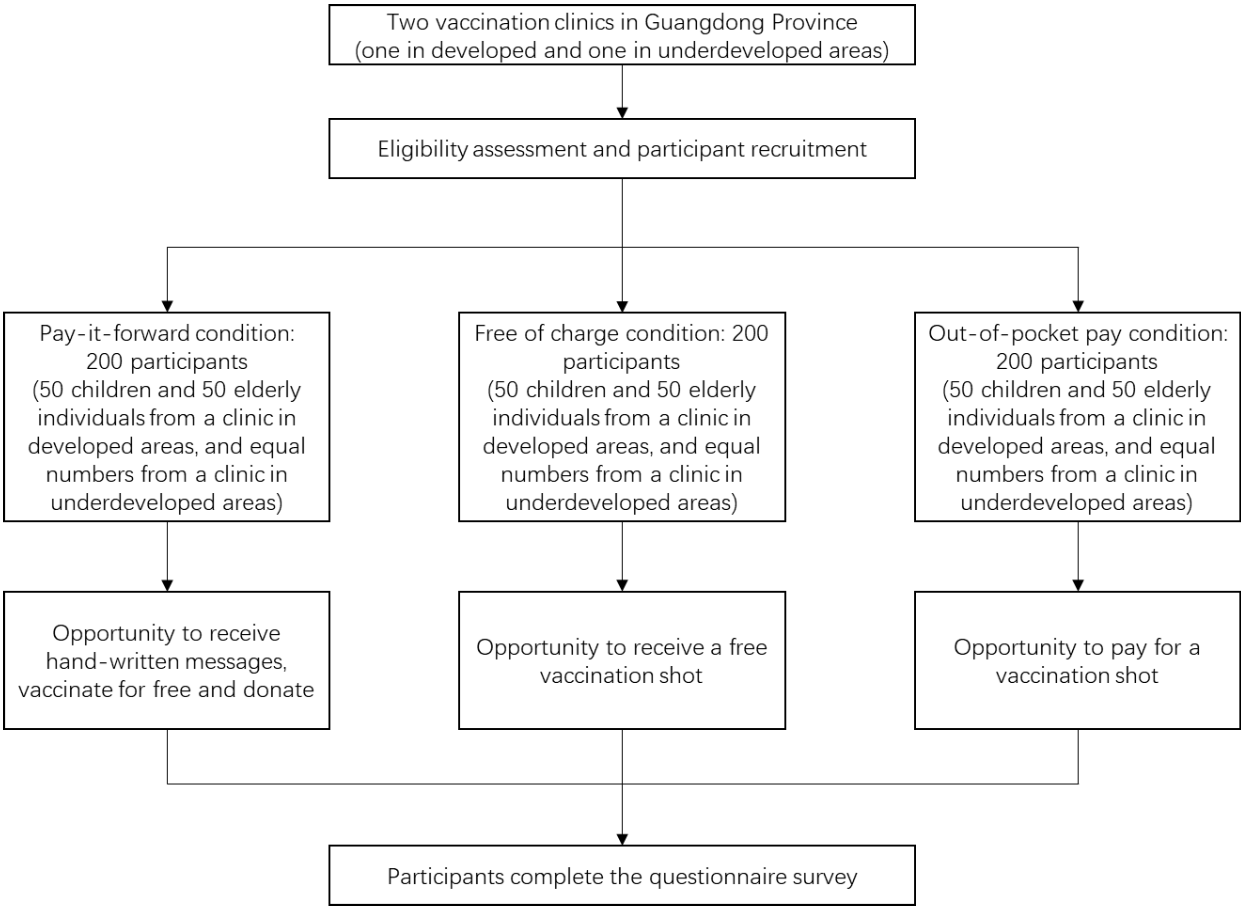


Figure.2

*Sampling and recruitment*

The target population will be recruited in selected vaccination clinics or communityhealth centers. Project staff will assess eligibility based on inclusion criteria as aforementioned and provide educational information about seasonal influenza pandemics to all potential participants, ie, guardians of children and older people. Participants will be recruited into three conditions in chronological order, namely pay-it-forward condition,free-of-charge condition, and standard care (out-of-pocket payment) condition.

Participants recruited during the period of pay-it-forward condition will be informed about the pay-it-forward vaccination program with handwritten warm messages co-created by participating families to enhance public trust and community cohesion (Supplementary files 1 and 2). After introducing the 'pay-it-forward' program, the projectstaff will provide eligible participants with the opportunity to take part, which means one eligible child or older individual can receive an influenza vaccination shot. The guardian or participant will then be asked whether they are willing to donate any amount of money into the pay-it-forward seed fund to support more future users to receive the same vaccination service. A donation collection box will be provided on site for those who prefer to donate cash and a WeChat QR code (a multifunctional social mobile app embedded with monetary transaction functions) will be provided to those who prefer online donation to the program. Neither the wiliness to donate nor the amount of donationwill affect them receiving the influenza vaccination services for free. The microdonationswill be used to support future users and usage will be publicized periodically in the vaccination clinic. Expected sample sizes for both children and older individuals in pay-it-forward condition are 100 respectively (50 children and 50 older individuals from the clinic in developed areas and equal numbers from the clinic in underdeveloped areas).

Participants recruited in the free-of-charge condition will be invited and provided a free influenza vaccination shot but they will not receive any community created messages about the pay-it-forward program. Participants recruited in the standard care condition will be asked to pay out of pocket in the event of being willing to receive the influenza vaccination shot. Participation in each condition is fully voluntary and anonymous. Access to other vaccination and medical services in the clinic will not be affected in any way due to the project. All participants will complete a short questionnaire survey to collect information about demographic backgrounds, information about participating pay-it-forward program (pay-it-forward condition only), and confidence and hesitancy in influenza vaccines.

*Data collection*

Written consent form will be obtained from all participants. This study will collect the following information: data recorded by the project staff using standard information tracking sheet including the number of invited and participating individuals, the number of participants who receive a vaccination shot, the number of individuals who donate and donation amount in the pay-it-forward condition, as well as survey data through the self-administered questionnaire. Those who have difficulty in reading the questionnaire survey will be assisted by the project staff on-site. A small gratitude gift worth of around 1 GBP will be given to each participant after completing the questionnaire survey.

*Risk assessment and management*

According to the World Health Organization reports, influenza vaccines are well tolerated,and side effects are mild and transient in general. Common adverse events include soreness, redness, and/or swelling at the vaccination site, fever, muscle pain, headache, nausea which do not require medical attention.[12] Severe threatening allergic reaction after vaccination, such as difficulty breathing, hoarseness or wheezing, swelling around the eyes or lips, hives, paleness, weakness, or a fast heart beat or dizziness, are rare. These signs most likely happen within a few minutes to a few hours after the vaccine is given. Participants who receive a vaccination shot will be required to stay at least half an hour in the vaccination clinic to be closely monitored by a designated nurse. They will be informed to self-monitor signs mentioned above for a few more hours after they leave the clinic. The contact number to a designated physician from the selected clinic will be provided to participants as emergency contact in the event of any severe adverse effectswithin 24 hours after the vaccination.

*Withdrawal of participants*

Participants can withdraw from the study at any time without giving any explanations.

Withdrawal from the study would not affect the participant's medical care or otherpreventive services at the clinic in any way.

*Reporting of any adverse events*

The local CDCs and vaccination clinics have established a strong surveillance system to monitor any adverse events associated with vaccinating patients. Each patient who receives an influenza vaccine is recorded in the clinical health information system and uploaded to the local CDCs' surveillance system. Patients with any adverse events will bedealt with by local health staff on a timely manner. The research team will leverage their existing platform for reporting any adverse events and keep a tracking record of adverse events. The research team will periodically meet with the designated health workers to make sure adverse events are properly treated.

*Data management*

Data will be managed according to the principles in accordance with good research practice specified by LSHTM Research Data Management Policy.[13] All data will be kept confidential and accessible only to trained study staff. All consent forms, administrative and survey data will be transformed into digital format and data in non-digital formatswill be stored in a securely locked cabinet at local institutes (Guangdong Second Provincial General Hospital in Guangzhou). Paper questionnaires will be filed and stored for at least 10 years after project completion as per LSHTM's Records Retention and Disposal Schedule. Study databases will be managed by a dedicated data manager (DM). All data will be checked and cleaned before being updated to the main database. The databases will be regularly backed up on the local server where back-ups are maintained in the disaster recovery room. Copies of the databases will be transferred to LSHTM using the fully encrypted (SSL) server.

The main risks to data security relate to loss of electronic data and breach of participant confidentiality. With the acquisition of a dedicated office, all data on computers will be stored in this office which will be locked overnight. Only data entry personnel and study personnel will have access to any of the electronic data. After all checks have been completed, data will be entered into databases using desktops. The data manager will upload double entered data at the end of each day onto their computer. The Data Managerwill create weekly back-ups on an external password protected hard drive. When necessary or required, and copies of this data will be securely transferred to LSHTMusing encrypted files. The risks for confidential breaching are minimal since most data will use anonymized IDs unique to each participant. We will ensure that all copies of the quantitative data files are password protected, with access to the password restricted to staff who need to work with the data. Also, we will report study results only at the aggregate level. To further minimize the risk of breaching confidentiality, all study staff will be trained and certified as part of MENISCUS-1 using the NIH online training course in Protecting Human Subject Research Participants and will be required to sign a staff confidentiality agreement form.

**Project Outcomes**

The main anticipated measurable outcomes are:

1) Influenza vaccination rate in each condition: vaccination rate of each condition will be calculated respectively through (dividing number of participants who are vaccinated by the total number of individuals invited to participate) * 100%. Statistical differences in vaccination rates of all three conditions will be examined.

2) Donation rate in the pay-it-forward condition: the donation rate of the pay-it-forward condition will be calculated through (dividing the number of participants who donate by the total number of

participants who receive a vaccination shot in the pay-it-forward group) * 100%

3) Donation measurements in the pay-it-forward condition: the total donation amount as well as proportion of economic costs covered by the microdonations through dividing total donation by total economic costs of vaccines used in the pay-it-forward condition.

**Data analysis**

Descriptive analyses of demographic and behavioral characteristics, participation rate, vaccination rate in each condition will be conducted. We will compare the vaccination rate between the pay-it-forward model, free-of-charge, and the standard of care model using chi-squared test and logistic regression, reported as crude odds ratios (cOR) and adjusted odds ratios (aOR), by adjusting for education, employment status, and urban/rural residence. We choose these covariates because they are potential confounders.We will use descriptive statistics to report the participants' awareness, confidence and hesitancy in influenza vaccines, contributions to future participants in the pay-it-forward group, and the fraction of the total cost of vaccination covered by pay-it-forward. All data will be analysed using SPSS Version 25.

**Audits and inspections**

The study may be subject audit by the London School of Hygiene & Tropical Medicine under their remit as sponsor, the Study Coordination Centre and other regulatory bodiesto ensure adherence to GCP.

**Insurance provision**

We will purchase around 10,000 USD insurance through the LSHTM insurance team as per LSHTM sponsorship and insurance policy.

**DMC/DSMB**

Given the low risks associated with a quasi-experimental study to improve existing publichealth service uptake, we believe having the project team as well as designate a data manager to meet periodically and monitor the data safety would suffice. DSMB will notbe assembled in this particular project.

**Ethics**

We have submitted local ethics application to Guangdong Second Provincial General Hospital and is currently under review. The LSHTM ethics committee will review the project protocol.

**Publication – section on who will write, named authors**

With inputs from the entire project team including LSHTM research team and local research team in China, the project manager Dan Wu and PI Joseph Tucker will lead the manuscript writing. More specifically, potential authors will include Dan Wu, Weiming Tang, Weibin Cheng, Yafei Si, Huipeng Liao, Yewei Xie, Nina Ren, Yunhui Chen, Zhangjun Tian, Joseph Tucker

**Confidentiality**

Each participant will be assigned a unique ID and personal identifiable information will not be collected. Participant survey data will only be used for research purpose of this particular project. Participant survey data will be transformed into digital format and copies of the quantitative data files will be password protected. Only designated project staff will have access to this encrypted information. We will report study results only at the aggregate level which will be difficult for others to identify individual participants. Without participant permission, project team researchers will not share the information toa third party.

10. Shu Y-L, Fang L-Q, de Vlas SJ, Gao Y, Richardus JH, Cao W-C. Dual seasonal patterns for

influenza, China. *Emerging infectious diseases* 2010; **16**(4): 725-6.

11. Xueqiao W, Tom H. China pharma crackdown leads to flu vaccine shortage. 2018. https://www.ft.com/content/6829cd0e-f07b-11e8-ae55-df4bf40f9d0d (accessed 19 Jan 2020).

12. WHO. Information sheet: Observed rate of vaccine reactions - Influenza vaccine. . 2012. https://www.who.int/vaccine_safety/initiative/tools/Influenza_Vaccine_rates_information_sheet.pdf (accessed 19 Jan 2020).

13. LSHTM. Research data management policy. 2019. https://www.lshtm.ac.uk/sites/default/files/research_data_management_policy.pdf (accessed 19 Jan 2020).

Supplementary file 1: Pay-it-forward

Research Proposal Version No.: 1

Research Proposal Version Date: Jan 28, 2020

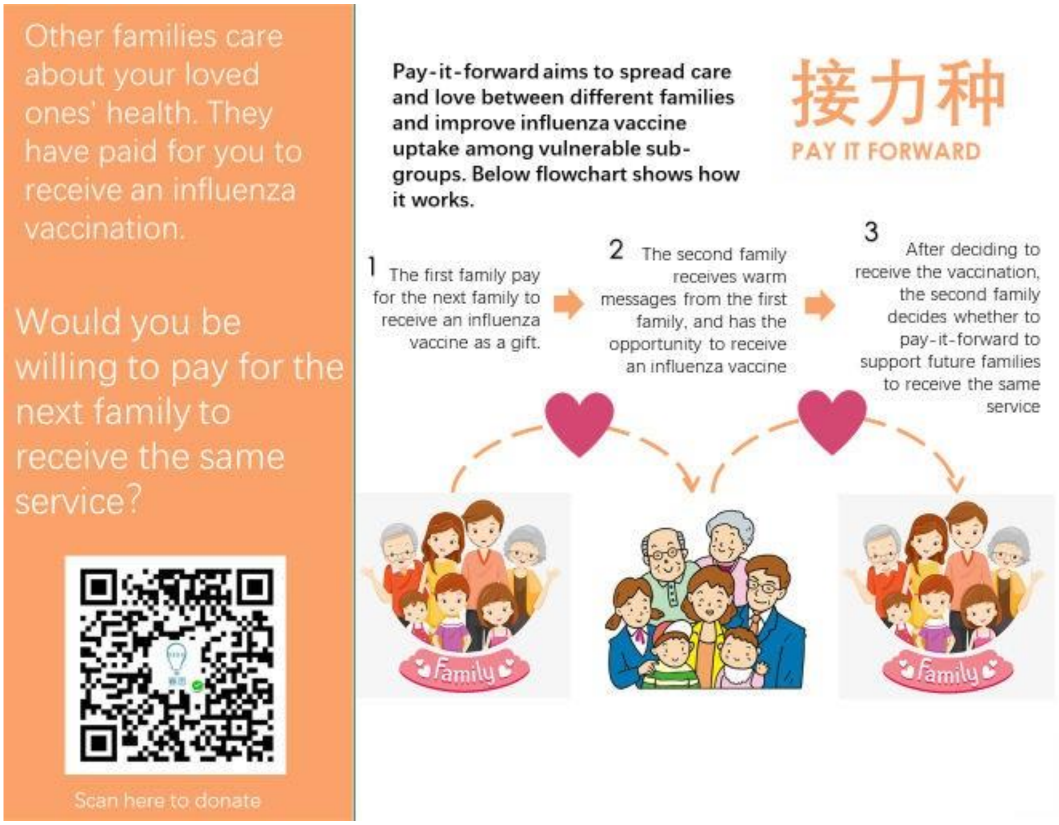


Supplementary file 2: Examples of hand-written messages

Research Proposal Version No.: 1

Research Proposal Version Date: Jan 28, 2020

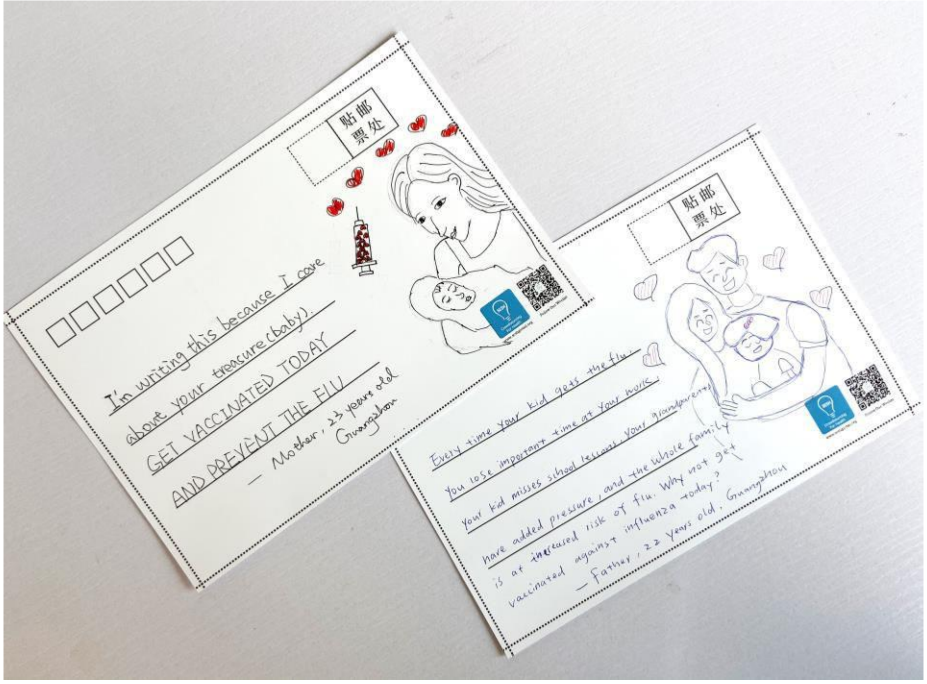

Research Proposal Version No.: 2

Research Proposal Version Date: April 14, 2020

**Title: Pay-it-forward to improve influenza vaccine uptake among children andolder people in China: A quasi-experimental study**

**Trial Sponsor**

Name: London School of Hygiene & Tropical Medicine

Address: London School of Hygiene & Tropical Medicine, Keppel Street, London WC1E7HT

**Project Summary**

Despite a high burden of influenza-related mortality, China has extremely low rates of influenza vaccination among key groups including children and older people. Ten people die of influenza each hour in China. Individuals ≥60 years is 26 times more like to die of influenza than people < 60. Kids (≤5 years) also have extremely high influenza related death. However, only 2% of Chinese are vaccinated. Influenza vaccines cost 9-16 GBP and are not covered by national insurance programs. We propose a pay-it-forward intervention to increase influenza vaccination rates among children aged between 6months and 8 years, as well as older people aged 60 or above in community-based health organizations. Pay-it-forward has one person receive a gift (e.g., a free influenza vaccination to a caregiver of a child or an old individual) and then asks them if theywould like to pay-it-forward for other individuals to benefit from the same generous gift. Participants will also be able to create handwritten postcards encouraging people in the local community to pay-it-forward to the next individual and donate to the rolling finance pool. We will evaluate the intervention using a quasi-experimental study at three community health organizations in Guangdong province, Southern China – one from developed Guangzhou city, one from under-developed Yangshan county, and one from Shenzhen city where free influenza vaccination is provided to children and older people. The primary comparison will be between pay-it-forward and standard of care conditions. We will also include a free vaccine treatment as an exploratory arm. We will recruit 50 children and 50 older individuals respectively during the pay-it-forward and standard of care treatment periods (25 children and 25 older individuals from each community vaccination clinic). We will recruit 50 children and 50 older individuals in free vaccine group in Shenzhen. In summary, a total of 300 participants will be recruited, including 150 children (consented by the guardian) and 150 older individuals. Primary outcomes are influenza vaccine uptake rates and pay-it-forward donation rate.

**Background**

Influenza is a viral respiratory infectious disease of global importance. Seasonal influenza epidemics cause 3 to 5 million severe cases and 290,000 to 650,000 deaths in the world each year.[1] Pregnant women, infants and children, older people, and patients with chronic diseases are at high risks of serious illness and death when infected. In China, about 10 people die from influenza-related illnesses each hour. [2] People older than 60 years old are26 times more like to die from influenza than younger individuals.[2] Influenza vaccinationis considered the most effective way to prevent influenza-related diseases. For example, globally, over 40% of countries have decided to provide free flu vaccine to key populations such as children and older individuals.

Research Proposal Version No.: 2

Research Proposal Version Date: April 14, 2020

Chinese Center for Diseases Control and Prevention (China CDC) has released in 2018 a high-level guideline for influenza vaccination services provision to at-risk populations and provinces are developing piloting programs. The guideline defines several at-risk populations including pregnant women, children, family members and caregivers ofinfants less than 6 months of age, and people aged over 60 years old. However, the most recent pooled evidence revealed that only 11.9% of children (aged 6 months to 17 years

old) and 21.7% of older population (aged >=60 years) were vaccinated in China.[3]

There are several reasons for low influenza vaccination rates in the country. First, influenza vaccines are not subsidized by government funding in most places due to governmental financial constraints and people need to pay around 16 pounds out-of- pocket for getting the vaccination. Second, the lack of trust in vaccines generally likely decreases demand for influenza vaccine in China. In July 2018, a massive vaccinescandal swept across China, and over 500,000 children were likely injected with faulty triple vaccines (diphtheria/pertussis/tetanus, DPT).[4] This scandal dramatically decreased trust towards vaccine programs, including influenza vaccine. In addition, pregnancy is widely considered a contraindication for influenza vaccination, and pregnant women in many places are rejected by health workers for vaccination.[5] In places where influenza vaccines are free, public awareness and vaccination rates are suboptimal compared to other countries.[6] The vast majority of the population are unaware of the vaccination and there is widespread hesitancy in safety and effectiveness of influenza vaccine, even among health

workers.[7] Only 8% of health workers always recommended vaccine to patients during the influenza season.[7] Innovative strategies are needed to reduce financial burden, build up public trust and collect evidence on effective interventions to improve vaccine uptake.

Research Proposal Version No.：2

Research Proposal Version Date：April 14, 2020

Pay-it-forward is an innovative, participatory behavioral intervention. Pay-it-forward has one individual receive a free influenza vaccine who will be informed by using hand- written postcard messages co-created by participants that someone else paid for them, then ask if they would like to support a future vaccination. Figure 1 illustrates how the model works. Our previous pay-it-forward studies focused on diagnostic services uptake among marginalized populations have proven effective in increasing chlamydia and gonorrhea tests uptake by three folds, compared to the control group.[8] In addition,89% of participants donated to the rolling finance pool and the community generosity build up trust in health services.[8] Evidence also showed simple acts of kindness are contagious [9]

and hold potential to be leveraged to enhance public health services delivery.

In this quasi-experimental study, we propose a pay-it-forward approach, aiming to improve public trust and influenza vaccine uptake among two key sub-groups, namely children aged between 6 months and 8 years old and older people aged 60 or above in Guangdong province. We determined these two sub-groups based on the disease burden and the national definition of at-risk populations. Specific objectives include 1) to examine the feasibility of applying the pay-it-forward approach in influenza vaccine research in the community setting; 2) to compare the effectiveness of three strategies, which are pay-it-forward, free of charge, and pay out-of-pocket (standard care condition), on influenza vaccine uptake among children and older individuals in China; 3) to investigate the donation rate and average donation amount among participants in the pay- it-forward condition.

Research Proposal Version No.: 2

Research Proposal Version Date: April 14, 2020

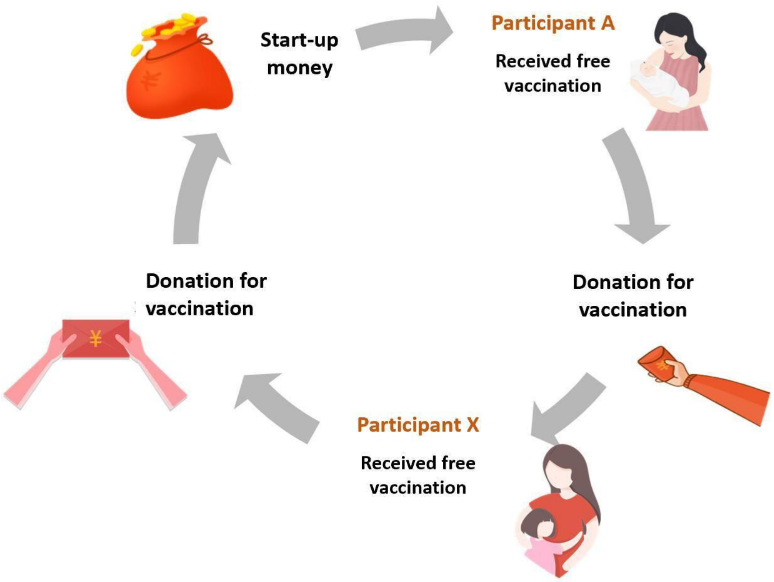


**Figure 1: "Pay-it-forward" Model**

**Hypotheses**

Based on the previous research, we conservatively estimate that compared to the standardcare (out-of-pocket pay) group,

1) "pay-it-forward" model can significantly improve the influenza vaccination rate among at-risk population including children and older groups (30% higher than the standard care group)
2) The donation rate of participants in the "pay it forward" group can reach 50% or higher
3) The "pay it forward" donation can cover 10% of the total economic cost of influenza vaccine.

**Methods**

Research Proposal Version No.: 2

Research Proposal Version Date: April 14, 2020

*Study setting*

Guangdong is a subtropical southern province with a population over 100 million in China. In southern China, influenza is prevalent throughout the year; it has a clear peak inthe summer and a less pronounced peak in the winter, including Guangdong.[10] In response to the national guideline to provide influenza vaccine to at-risk populations, Guangdong Province has released relevant provincial level policies to promote influenza vaccine. Stockouts of influenza vaccines have been reported in different regions due to tighter quality control of pharmaceutical products after the national vaccine scandal in 2018.[11] Therefore, identifying study sites with sufficient vaccine supply is key to the success of the pilot study during the post-scandal transitioning period. Two batches of tetravalent influenza vaccines produced by Hualan Biology company have been issued in August, 2019; and the first 500,000 tetravalent influenza vaccines were supplied to eight

provinces in China, including Guangdong. Given above factors, Guangdong province is selected as the major study location. The London School of Hygiene and Tropical Medicine (LSHTM) research team has established a local collaborative institute with strong primary care networks in the province - Guangdong Second Provincial General Hospital.

*Target population and inclusion criteria*

The pilot study will focus on children and older people. The inclusion criteria of this study are divided into two age groups:

1） Children: i) children aged between six-month-old and eight years old, ii) no acute moderate or severe illnesses with or without fever, iii) no severe, life-threatening allergies to flu vaccine or any ingredient(s) in the vaccine., iv) a legal guardian, be it a parent or grandparent in the Chinese setting, consents to participate the study.

2） Older people: i) ≥ sixty years old, ii) no acute moderate or severe illnesses with or without fever, iii) no severe, life-threatening allergies to flu vaccine or any ingredient(s) in the vaccine, iv) intelligently capable of making informed decisions
and consent to participate the study.

*Study design*

In this study, three vaccination clinics – one in developed Guangzhou city, one inunderdeveloped Yangshan county, and one in Shenzhen city where free influenza vaccines are provided to children and older people – will be selected in Guangdong Province for recruitment. The selected community health centers or vaccination clinics should have established infrastructure to provide influenza vaccination services, e.g. have sufficient influenza vaccine stock, relevant medical personnel including designated physicians and nurses for providing vaccine services. The research design is shown in Figure 2.

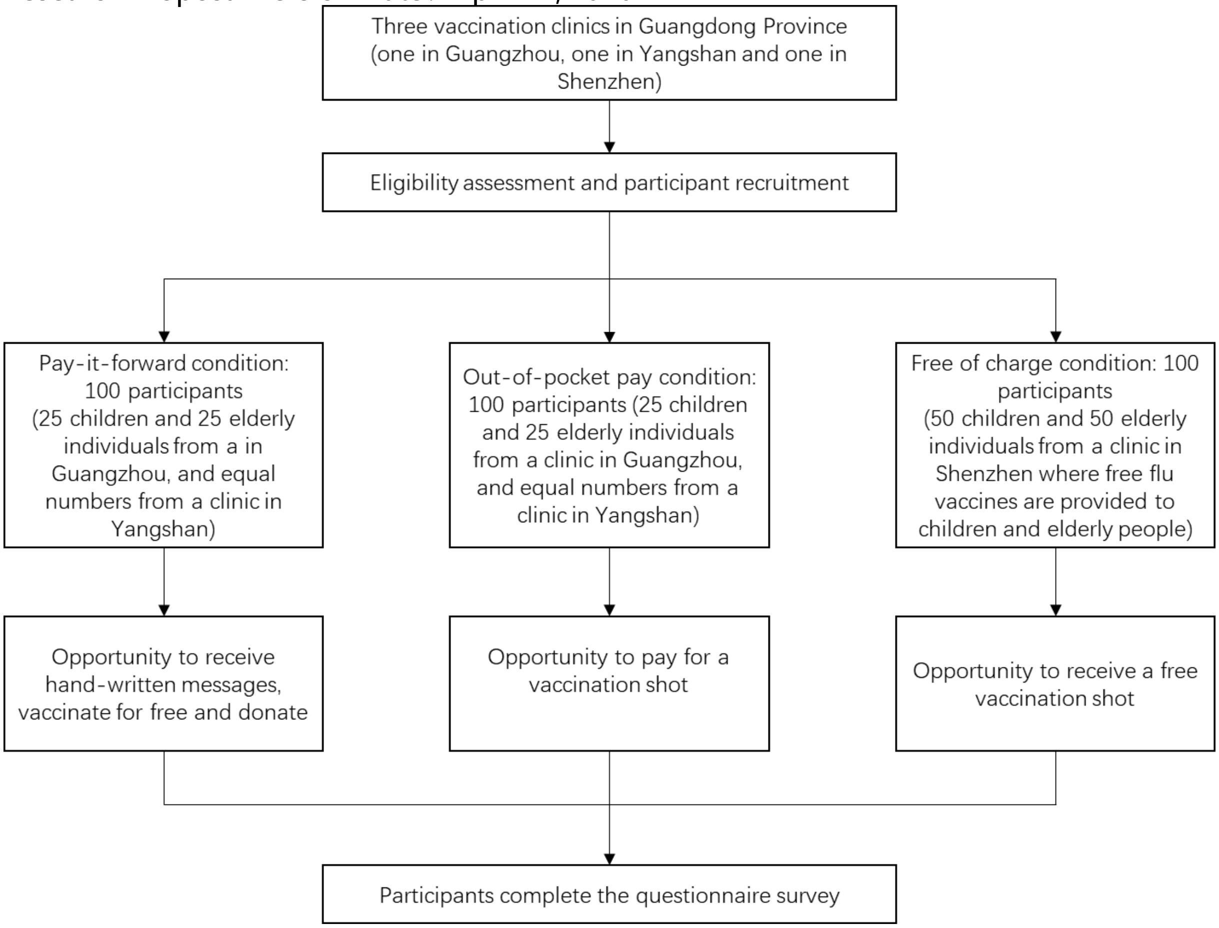


Figure.2

Sample size

Given the differences between children and older adults, we stratified sample size calculations by age groups. Based on our pilot data, we optimistically estimated the vaccine uptake as 30% in the standard-of-care condition, and conservatively estimated the vaccine uptake in the pay-it-forward condition as 80%, and a significance level of 0.025. Thus, a sample size of 100 (50 in the control and 50 in the pay-it-forward condition) for each age group would give us 90% power to test that the pay-it-forward is superior to the stand of care in promoting vaccination uptake, with a margin of 10%. We will include a free vaccine condition that provides free vaccines without any community engagement. We include this additional arm because this provides an opportunity to compare the cost of pay-it-forward and free service provision. However, the study is not powered to assess the difference between pay-it-forward and free arms. Given the primary comparison will be between pay-it-forward and standard of care arms, we will therefore implement the free vaccine arm as an exploratory arm (n = 100). In summary, the total estimated samples sizes are 150 children and 150 older people for the study (50 participants for each condition by age group).

*Sampling and recruitment*

The target population will be recruited in selected vaccination clinics or communityhealth centers. Project staff will assess eligibility based on inclusion criteria as aforementioned and provide educational information about seasonal influenza to all potential participants, ie, guardians of children and older people. Participants will be recruited into three conditions, namely pay-it-forward condition and standard care (out- of-pocket payment) condition at the Guangzhou and Yangshan sites, and free-of-charge condition in Shenzhen study site.

Participants recruited during the period of pay-it-forward condition will be informed about the pay-it-forward vaccination program with handwritten warm messages co- created by participating families to enhance public trust and community cohesion (Supplementary files 1 and 2). After introducing the ‘pay-it-forward’ program, the projectstaff will provide eligible participants with the opportunity to take part, which means one eligible child or older individual can receive an influenza vaccination shot. The guardian or participant will then be asked whether they are willing to donate any amount of money into the pay-it-forward seed fund to support more future users to receive the same vaccination service. A donation collection box will be provided on site for those who prefer to donate cash and a WeChat QR code (a multifunctional social mobile app embedded with monetary transaction functions) will be provided to those who prefer online donation to the program. Neither the willingness to donate nor the amount of donation will affect them receiving the influenza vaccination services for free. Themicrodonations will be used to support future users and usage will be publicized periodically in the vaccination clinic. Expected sample sizes for both children and olderindividuals in pay-it-forward condition are 50 respectively (25 children and 25 older individuals from the clinic in Guangzhou and equal numbers from the clinic in Yangshan) (Figure 2).

Participants for the free-of-charge condition will be recruited at a clinic in Shenzhen.They will be invited and provided a free influenza vaccination shot but they will not receive any community created messages about the pay-it-forward program. Expected sample sizes for both children and older individuals in free-of-charge condition are 50 respectively (Figure 2). Participants recruited in the standard care condition will be asked to pay out of pocket in the event of being willing to receive the influenza vaccination shot.Expected sample sizes for both children and older individuals in standard-of-care condition are 50 respectively (25 children and 25 older individuals from the clinic in Guangzhou and equal numbers from the clinic in Yangshan) (Figure 2). Participation in each condition is fully voluntary and anonymous. Access to other vaccination and medical services in the clinic will not be affected in any way due to the project. All participants will complete a short questionnaire survey to collect information about demographic backgrounds, information about participating pay-it-forward program (pay- it-forward condition only), and confidence and hesitancy in influenza vaccines.

*Data collection*

Written consent form will be obtained from all participants. This study will collect the following information: data recorded by the project staff using standard information tracking sheet including the number of invited and participating individuals, the number of participants who receive a vaccination shot, the number of individuals who donate and donation amount in the pay-it-forward condition, as well as survey data through the self- administered questionnaire. Those who have difficulty in reading the questionnaire survey will be assisted by the project staff on-site. A small gratitude gift worth of around 1 GBP will be given to each participant after completing the questionnaire survey.

*Risk assessment and management*

According to the World Health Organization reports, influenza vaccines are well tolerated,and side effects are mild and transient in general. Common adverse events include soreness, redness, and/or swelling at the vaccination site, fever, muscle pain, headache, nausea which do not require medical attention.[12] Severe threatening allergic reaction after vaccination, such as difficulty breathing, hoarseness or wheezing, swelling around the eyes or lips, hives, paleness, weakness, or a fast heart beat or dizziness, are rare. These signs most likely happen within a few minutes to a few hours after the vaccine is given. Participants who receive a vaccination shot will be required to stay at least half an hour in the vaccination clinic to be closely monitored by a designated nurse. They will be informed to self-monitor signs mentioned above for a few more hours after they leave the clinic. The contact number to a designated physician from the selected clinic will be provided to participants as emergency contact in the event of any severe adverse effectswithin 24 hours after the vaccination.

*Withdrawal of participants*

Participants can withdraw from the study at any time without giving any explanations. Withdrawal from the study would not affect the participant's medical care or other

preventive services at the clinic in any way.

*Reporting of any adverse events*

The local CDCs and vaccination clinics have established a strong surveillance system to monitor any adverse events associated with vaccinating patients. Each patient who receives an influenza vaccine is recorded in the clinical health information system and uploaded to the local CDCs' surveillance system. Patients with any adverse events will bedealt with by local health staff on a timely manner. The research team will leverage their existing platform for reporting any adverse events and keep a tracking record of adverse events. The research team will periodically meet with the designated health workers to make sure adverse events are properly treated.

*Data management*

Data will be managed according to the principles in accordance with good research practice specified by LSHTM Research Data Management Policy.[13] All data will be kept confidential and accessible only to trained study staff. All consent forms, administrative and survey data will be transformed into digital format and data in non-digital formatswill be stored in a securely locked cabinet at local institutes (Guangdong Second Provincial General Hospital in Guangzhou). Paper questionnaires will be filed and stored for at least 10 years after project completion as per LSHTM's Records Retention and Disposal Schedule. Study databases will be m

anaged by a dedicated data manager (DM). All data will be checked and cleaned before being updated to the main database. The databases will be regularly backed up on the local server where back-ups are maintained in the disaster recovery room. Copies of the databases will be transferred to LSHTM using the fully encrypted (SSL) server.

The main risks to data security relate to loss of electronic data and breach of participant confidentiality. With the acquisition of a dedicated office, all data on computers will be stored in this office which will be locked overnight. Only data entry personnel and study personnel will have access to any of the electronic data. After all checks have been completed, data will be entered into databases using desktops. The data manager will upload double entered data at the end of each day onto their computer. The Data Managerwill create weekly back-ups on an external password protected hard drive. When necessary or required, and copies of this data will be securely transferred to LSHTMusing encrypted files. The risks for confidential breaching are minimal since most data will use anonymized IDs unique to each participant. We will ensure that all copies of the quantitative data files are password protected, with access to the password restricted to staff who need to work with the data. Also, we will report study results only at the aggregate level. To further minimize the risk of breaching confidentiality, all study staff will be trained and certified as part of MENISCUS-1 using the NIH online training course in Protecting Human Subject Research Participants and will be required to sign a staff confidentiality agreement form.

**Project Outcomes**

The main anticipated measurable outcomes are:

1) Influenza vaccination rate in each condition: vaccination rate of each condition will be calculated respectively through (dividing number of participants who are vaccinated by the total number of individuals invited to participate) * 100%. Statistical differences in vaccination rates of all three conditions will be examined.

2) Donation rate in the pay-it-forward condition: the donation rate of the pay-it-forward condition will be calculated through (dividing the number of participants who donate by the total number of participants who receive a vaccination shot in the pay-it-forward group) * 100%

3) Donation measurements in the pay-it-forward condition: the total donation amount as well as proportion of economic costs covered by the microdonations through dividing total donation by total economic costs of vaccines used in the pay-it-forward condition.

**Data analysis**

Descriptive analyses of demographic and behavioral characteristics, participation rate, vaccination rate in each condition will be conducted. We will compare the vaccination rate between the pay-it-forward model, free-of-charge, and the standard of care model using chi-squared test and logistic regression, reported as crude odds ratios (cOR) and adjusted odds ratios (aOR), by adjusting for education, employment status, and urban/rural residence. We choose these covariates because they are potential confounders.We will use descriptive statistics to report the participants' awareness, confidence and hesitancy in influenza vaccines, contributions to future participants in the pay-it-forward group, and the fraction of the total cost of vaccination covered by pay-it-forward. All data will be analysed using SPSS Version 25.

**Audits and inspections**

The study may be subject audit by the London School of Hygiene & Tropical Medicine under their remit as sponsor, the Study Coordination Centre and other regulatory bodies to ensure adherence to GCP.

**Insurance provision**

We will purchase around 10,000 USD insurance through the LSHTM insurance team as per LSHTM sponsorship and insurance policy.

**DMC/DSMB**

Given the low risks associated with a quasi-experimental study to improve existing publichealth service uptake, we believe having the project team as well as designate a data manager to meet periodically and monitor the data safety would suffice. DSMB will not be assembled in this particular project.

**Ethics**

We have submitted local ethics application to Guangdong Second Provincial General Hospital and is currently under review. The LSHTM ethics committee will review the project protocol.

**Publication – section on who will write, named authors**

With inputs from the entire project team including LSHTM research team and local research team in China, the project manager Dan Wu and PI Joseph Tucker will lead the manuscript writing. More specifically, potential authors will include Dan Wu, Weiming Tang, Weibin Cheng, Yafei Si, Huipeng Liao, Yewei Xie, Nina Ren, Yunhui Chen, Zhangjun Tian, Joseph Tucker

**Confidentiality**

Each participant will be assigned a unique ID and personal identifiable information will not be collected. Participant survey data will only be used for research purpose of this particular project. Participant survey data will be transformed into digital format and copies of the quantitative data files will be password protected. Only designated project staff will have access to this encrypted information. We will report study results only at the aggregate level which will be difficult for others to identify individual participants. Without participant permission, project team researchers will not share the information toa third party.

10. Shu Y-L, Fang L-Q, de Vlas SJ, Gao Y, Richardus JH, Cao W-C. Dual seasonal patterns for

influenza, China. *Emerging infectious diseases* 2010; **16**(4): 725-6.

11. Xueqiao W, Tom H. China pharma crackdown leads to flu vaccine shortage. 2018. https://www.ft.com/content/6829cd0e-f07b-11e8-ae55-df4bf40f9d0d (accessed 19 Jan 2020).

12. WHO. Information sheet: Observed rate of vaccine reactions - Influenza vaccine. . 2012. https://www.who.int/vaccine_safety/initiative/tools/Influenza_Vaccine_rates_information_sheet.pdf (accessed 19 Jan 2020).

13. LSHTM. Research data management policy. 2019. https://www.lshtm.ac.uk/sites/default/files/research_data_management_policy.pdf (accessed 19 Jan 2020).

Supplementary file 1: Pay-it-forward

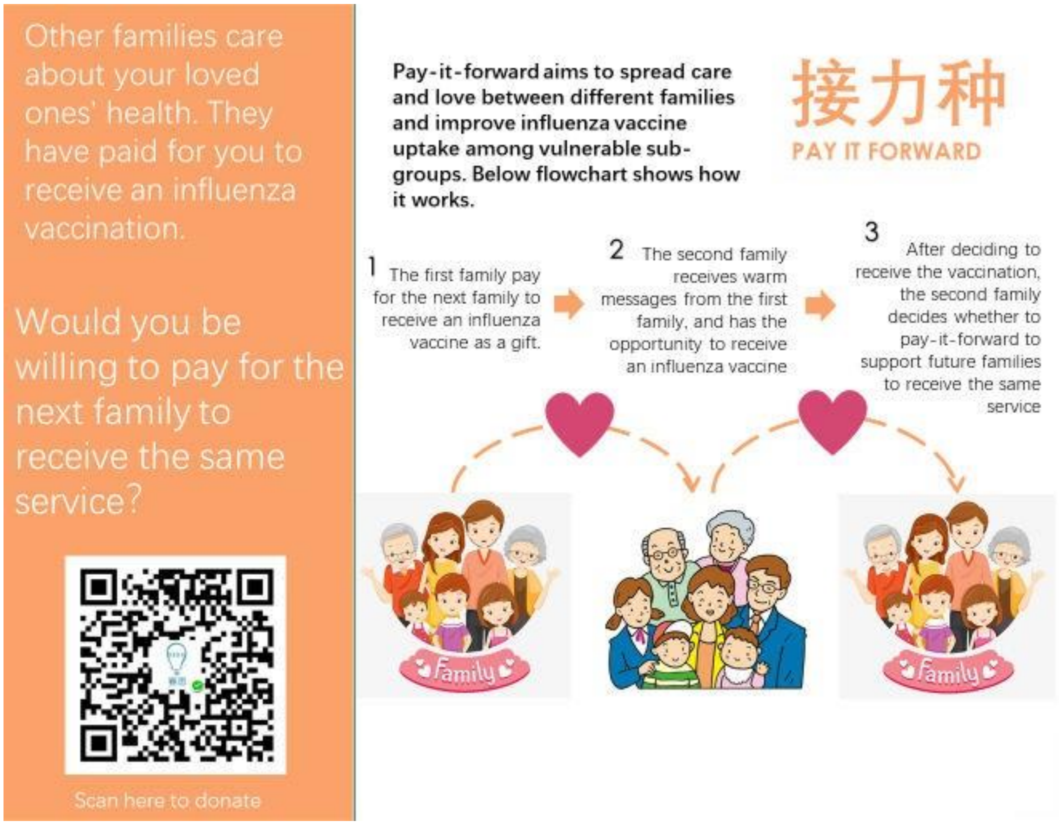

Research Proposal Version No.：2

Research Proposal Version Date：April 14, 2020

Supplementary file 2: Examples of hand-written messages

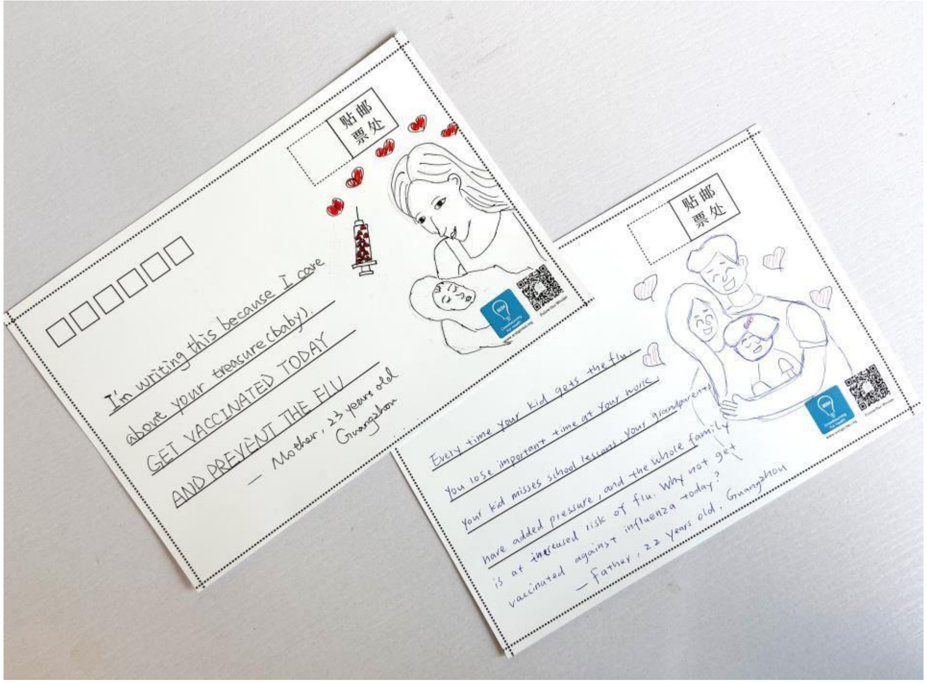

Research Proposal Version No.: 3

Research Proposal Version Date: Feb 9, 2021

**Title: Pay-it-forward to improve influenza vaccine uptake among children and olderadults in China: A three-arm quasi-experimental study**

**Trial Sponsor**

Name: London School of Hygiene & Tropical Medicine

Address: London School of Hygiene & Tropical Medicine, Keppel Street, London WC1E7HT

**Protocol Version 1.3 (February 9, 2021)**

**China Clinical Trials Number: ChiCTR2000040048****Project Summary**

Despite a high burden of influenza-related mortality, China has low rates of influenza vaccination among key groups including children and older people. Ten people die ofinfluenza each hour in China. However, only 2% of Chinese are vaccinated and mostpeople in China have limited understanding of influenza vaccines. Influenza vaccinescost 9-16 GBP and are not covered by the government or employers. We propose a pay-it-forward intervention to increase influenza vaccination uptake among children and older people in China. Pay-it-forward has one person receive a gift (e.g., a free influenza vaccination) and then asks them if they would like to pay-it-forward for other individualsto receive a free vaccine. Participants will also be able to create handwritten postcards encouraging people in the local community to pay-it-forward to the next individual and donate to the rolling finance pool.

We will evaluate the intervention using a quasi-experimental study at three community health centers in Guangdong province, Southern China – one from urban Guangzhou city, one from suburban Zengcheng city, and one from under-developed rural Yangshan county. The primary comparison will be between pay-it-forward and standard of care conditions. We will also include a free vaccine treatment as an exploratory arm. Three arms will be implemented at all three health centers. We will recruit 75 children and 75 older individuals respectively during the pay-it-forward, standard of care and free vaccine treatment periods (25 children and 25 older individuals from each community vaccination clinic). In summary, a total of 450 participants will be recruited, including 225 children (consented by the guardian) and 225 older individuals. Primary outcomes are influenza vaccine uptake rates and pay-it-forward donation rate.

**Background**

Influenza is a viral respiratory infectious disease of global importance. Seasonal influenzacause 3 to 5 million severe cases and 290,000 to 650,000 deaths in the world each year.[1] Pregnant women, infants and children, older people, and patients with chronic diseases are at high risks of serious illness and death when infected. In China, about 10 people die from influenza-related illnesses each hour. [2] People older than 60 years old are 26 times more likely to die from influenza than younger individuals.[2] Influenza vaccination is considered the most effective way to prevent influenza-related diseases. For example, globally, over 40% of countries have policies to provide free flu vaccine to keypopulations.

Chinese Center for Diseases Control and Prevention (China CDC) has released a high-level guideline for influenza vaccination services provision to at-risk populations in2018. The guideline defines several at-risk populations including pregnant women, children, family members and caregivers of infants less than 6 months of age, and peopleaged over 60 years old. However, the most recent pooled evidence by 2016 revealed thatonly 11.9% of children (aged 6 months to 17 years old) and 21.7% of older people (aged >=60 years) were vaccinated in China.[3]

There are several reasons for low influenza vaccination rates in the country. First, influenza vaccines are not subsidized by government funding in most places and people need to pay around 16 pounds out-of-pocket for getting the vaccination. Second, the lackof trust in vaccines due to a massive vaccine scandal in China is also considered to decrease demand for influenza vaccine in China.[4] In addition, pregnancy is widely considered a contraindication for influenza vaccination, and pregnant women in many places are rejected by health workers for vaccination.[5] Even in places where influenza vaccines are free, public awareness and vaccination rates are suboptimal compared to other countries.[6] The widespread hesitancy in safety and effectiveness of influenza vaccine would be the main reason for this phenomenon.[7] Innovative strategies are needed to reduce financial burden, build up public trust and collect evidence on effectiveinterventions to improve vaccine uptake.

Pay-it-forward is an innovative, participatory behavioral intervention. Pay-it-forward hasone individual receive a free influenza vaccine who will be informed by using hand-written postcard messages co-created by participants that someone else paid for them, then ask if they would like to support a future vaccination. Figure 1 illustrates howthe model works. Our previous pay-it-forward studies focused on diagnostic services uptake among marginalized populations have proven effective in increasing chlamydia and gonorrhea tests uptake by three folds, compared to the control group.[8] In addition,89%of participants donated to the rolling finance pool and the community generosity built up trust in health services.[8] Evidence also showed simple acts of kindness are contagious [9] and hold potential to be leveraged to enhance public health services delivery.

Research Proposal Version No.: 3

Research Proposal Version Date: Feb 9, 2021

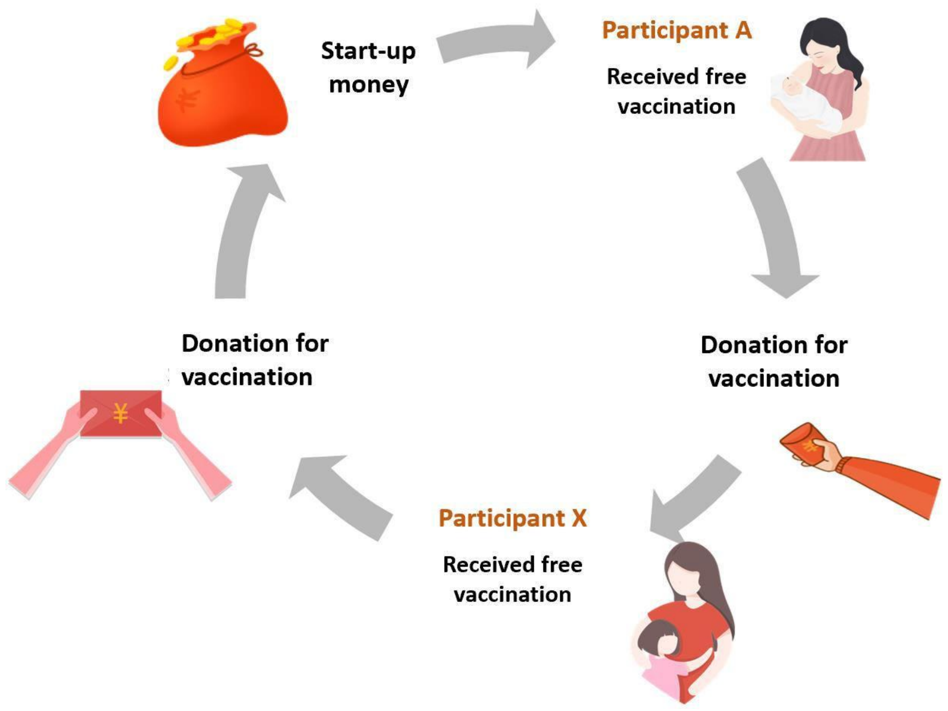


**Figure 1: "Pay-it-forward" Model**

In this quasi-experimental study, we propose a pay-it-forward approach, aiming to improve influenza vaccine uptake among two key sub-groups, namely children aged between 6 months and 8 years old and older adults aged 60 or above in Guangdong province of China. We determined these two sub-groups based on the disease burden, thenational definition of at-risk populations and local clinical practice guidelines. Specific objectives include 1) to examine the effectiveness of pay-it-forward approach on influenza vaccine uptake among Chinese people in comparison to standard-of-care and free vaccination; 2) to conduct an economic evaluation of pay-it-forward on vaccine uptake in comparison to free vaccination and standard-of-care; and 3) to examine factors associated with vaccine uptake, confidence/hesitancy, and other secondary outcomes.

### Hypotheses

Our main hypotheses include the following:

1) Pay-it-forward can significantly improve the influenza vaccination rate among at-risk populations compared to the standard-of-care arm
2) Pay-it-forward is more cost-effective than standard-of-care in terms of cost perperson vaccinated.
3) Many participants will donate money to support subsequent people to receive a freevaccine.
4) Influenza vaccine uptake in the pay-it-forward arm will be similar to that of the freevaccine arm (exploratory).

### Methods

*Study setting*

Guangdong is a subtropical southern province in China with a population over 100 million. In southern China, influenza is prevalent throughout the year with a clear peak inthe summer and a less pronounced peak in the winter.[10] In response to the national guideline to provide influenza vaccine to at-risk populations, Guangdong Province has released relevant provincial level policies to promote influenza vaccine. Stockouts of influenza vaccines have been reported in different regions due to tighter quality control ofpharmaceutical products after the national vaccine scandal in 2018.[11] We also liaised withSanofi China office and Guangdong Second Provincial General Hospital to help identify study sites that have sufficient influenza vaccines.

*Intervention development*

Our team co-created messages as part of a participatory hackathon in Tanzania and a nationwide crowdsourcing open call in China. First, our team worked with diverse individuals as part of a three-day

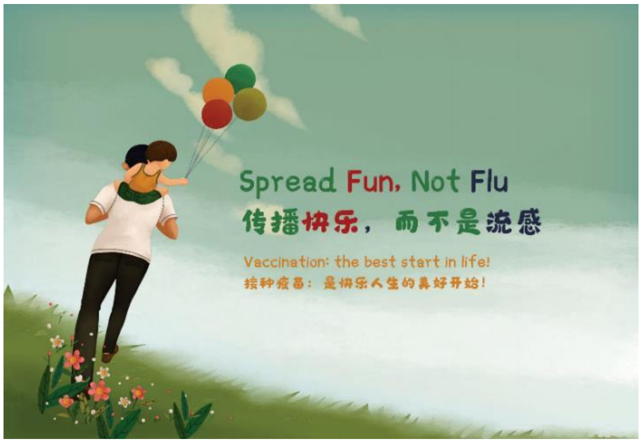


**Fig 2: Postcard example**

hackathon organized by the Bill and Melinda Gates Foundation and the World Food Program in Tanzania. We identified engagementstrategies, including 1) inviting participants to co-create postcard messages for the pre-test (see Fig 2 for an example); 2) organizing a nationwide crowdsourcing open callto engage public in generating health messages focused on influenza and vaccines; 3) engaging local community staff in implementing the pay-it-forward pragmatic trial.

Then, our partner SESH (Social Entrepreneurship to Spur Health) led a nationwide crowdsourcing open contest to solicit messages promoting influenza vaccination. The crowdsourcing open call had five steps – organizing a steering committee to oversee the process, promoting the open call via digital and in-person methods, evaluating submissions (texts, images, videos), celebrating exceptional submissions, and sharing andimplementing exceptional ideas. Due to COVID-19, the open call was transitioned to a mostly digital format. The open call website was viewed 7151 times according to analytics. We received a total of 305 submissions, including 204 texts, 90 images, and three videos. All eligible submissions were evaluated by a crowd judge panel and an expert judge panel based on pre-defined criteria (novelty, relevance, feasibility, and elaboration). Crowd judges were invited via social media platforms on a voluntary basis and each submission was judged by at least three independent crowd judges. Experts in public health research, health communication, vaccines, and infectious diseases were invited to the expert panel. Submissions were scored on a 1 to 10 scale and those ranked in the top 15% were deemed exceptional. Some of these messages were integrated into the recruitment pamphlet for the study.

*Study population and inclusion criteria*

The study will focus on children and older adults. The inclusion criteria of this study aredivided into two age groups:

1） Parents or guardians of children between six months and eight years old. Each childneeded to meet the following eligibility criteria: i) no acute moderate or severe illnesses, ii) eligible to receive an influenza vaccine based on clinical evaluation froma physician; iii) has a legal guardian (e.g. a parent or grandparent) who lives in Chinaand consents to participate in the study; and iv) has not received an influenza vaccinein the past year.

2） Older people: i) ≥ sixty years old; ii) no acute moderate or severe illness; iii) eligibleto receive an influenza vaccine based on clinical evaluation from a physician; iv) capable of making informed decisions and consenting to participate in the study; and v) have not received an influenza vaccine in the past year.

*Study design*

In this study, three sites –one in rural Yangshan county, one in the suburban Zengcheng district, and one in urban Guangzhou city will be selected in Guangdong Province for recruitment, to represent the three different economic conditions in Guangdong province.The selected community health centers or vaccination clinics should have established infrastructure to provide influenza vaccination services, e.g. have sufficient influenza vaccine stock, relevant medical personnel including designated physicians and nurses forproviding vaccine services. The research design is shown in Figure 3.

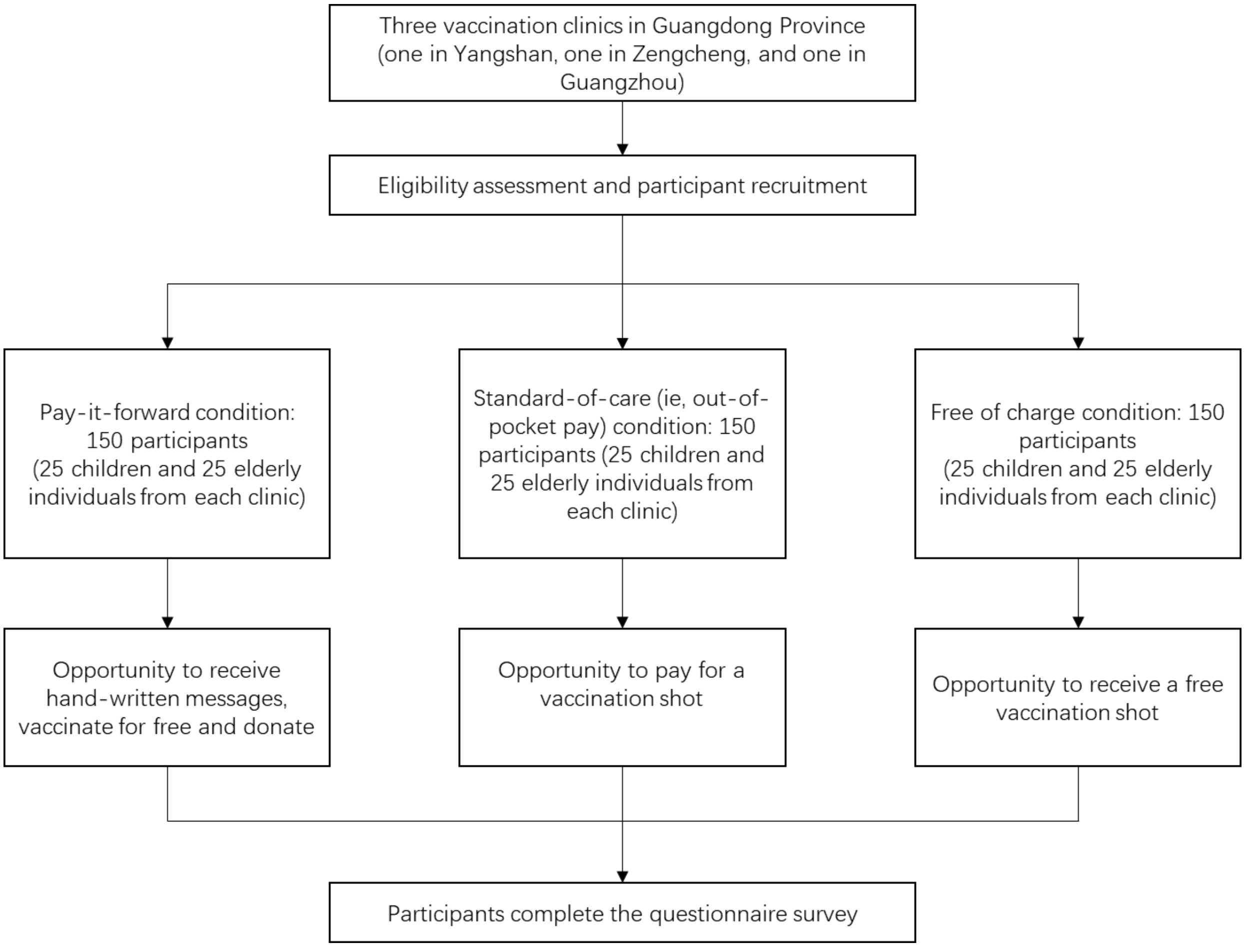


Figure 3: Flowchart of the quasi-experimental study

Sample size

Given the differences between children and older adults, we stratified sample size calculations by age groups. Based on our pilot data, we optimistically estimated the vaccine uptake as 30% in the standard-of-care condition, and conservatively estimated the vaccine uptake in the pay-it-forward condition as 80%, and a significance level of 0.025. Thus, a sample size of 100 (50 in the control and 50 in the pay-it-forward condition) for each age group would give us 90% power to test that the pay-it-forward is superior to the stand of care in promoting vaccination uptake, with a margin of 10%. We will include a free vaccine condition that provides free vaccines without any community engagement. We include this additional arm because this provides an opportunity to compare the cost of pay-it-forward and free service provision. However, the study is not powered to assess the difference between pay-it-forward and free arms. Given the primary comparison will be between pay-it-forward and standard of care arms, we will therefore implement the free vaccine arm as an exploratory arm.

To allow sub-analysis by age group, we will recruit 25 more children and older individuals respectively for each arm as a boost sample of each age group. In summary, 150 participants (75 children and 75 older adults) will be recruited for each arm, totaling 450 participants for the study.

*Sampling and recruitment*

The study participants will be recruited in selected vaccination clinics or community health centers. Project staff will assess eligibility based on inclusion criteria as aforementioned and provide educational information about seasonal influenza to all potential participants, ie, guardians of children and older people. Participants will berecruited into three arms, namely standard-of-care (out-of-pocket payment) arm,

pay-it-forward arm, and free-of-charge arm at all three sites on a time-based recruitmentmanner. In order to decrease the effect of temporal trends, we will minimize the time between participants receiving the standard-of-care treatment and participants receivingthe pay-it-forward treatment.

*Standard-of-care*: Participants recruited in the standard care arm will be asked to pay out

of pocket in the event of being willing to receive the influenza vaccination shot. Expectedsample sizes for both children and older adults in standard-of-care arm are 75 respectively (25 children and 25 older adults from each clinic) (Figure 3).

*Pay-it-forward Intervention*: Participants recruited during the period of pay-it-forward

arm will be informed about the pay-it-forward vaccination program with handwritten warm messages co-created by participating families to enhance public trust and community cohesion (Supplementary files 1 and 2). After introducing the 'pay-it-forward'program, the project staff will provide eligible participants with the opportunity to take part, which means one eligible child or older individual can receive an influenza vaccine. The guardian or participant will then be asked whether they are willing to donate any amount of money into the pay-it-forward seed fund to support more future users to receive the same vaccination service. A donation collection box will be provided on site for those who prefer to donate cash and a WeChat QR code (a multifunctional social mobile app embedded with monetary transaction functions) will be provided to those whoprefer online donation to the program. Neither the willingness to donate nor the amount of donation will affect them receiving the influenza vaccination services for free. The microdonations will be used to support future users and usage will be publicized periodically. Expected sample sizes for both children and older adults in pay-it-forward arm are 75 respectively (25 children and 25 older individuals from each clinic) (Figure 3).

*Free-of-charge*: Participants will be invited and provided a free influenza vaccine but

they will not receive any community created messages about the pay-it-forward program.Expected sample sizes for both children and older adults in free-of-charge arm are 75 respectively (25 children and 25 older individuals from each clinic) (Figure 3).

Participation in each arm is fully voluntary and anonymous. Access to other vaccinationand medical services in the clinic will not be affected in any way due to the project. All participants will complete a short questionnaire survey to collect information about demographic backgrounds, prior influenza vaccine history, information about participating pay-it-forward program (pay-it-forward arm only), and confidence and hesitancy in influenza vaccines.

*Data collection*

Written consent form will be obtained from all participants. This study will collect the following information: data recorded by the project staff using standard information tracking sheet including the number of invited and participating individuals, the number of participants who receive a vaccine, the number of individuals who donate and donationamount in the pay-it-forward arm, as well as survey data through the self-administered online questionnaire. Those who have difficulty in reading the questionnaire survey will be assisted by the project staff on-site. A small gratitude gift worth of around 1 GBP will be given to each participant after completing the questionnaire survey.

*Outcomes*

The primary outcome of the study will be influenza vaccine uptake. This will be assessed by administrative records. Secondary outcomes include costs outcomes, vaccine confidence, vaccine hesitancy, and adverse events by administrative records. The main comparison will be between pay-it-forward and standard-of-care arms. Outcomes will also be stratified by age group (children and older people) and study sites.

*Data analysis*

Descriptive analyses of demographic and behavioral characteristics, participation rate, vaccination rate in each arm will be conducted. We will compare the vaccination rate between standard of care, pay-it-forward, and free-of-charge arms using chi-squared testand logistic regression, reported as crude odds ratios (cOR) and adjusted odds ratios (aOR), by adjusting for education, employment status, and study sites. We choose these covariates because they are potential confounders. We will use descriptive statistics to report the participants' awareness, prior influenza vaccination history, confidence and hesitancy in influenza vaccines, contributions to future participants in the pay-it-forwardarm, and the fraction of the total cost of vaccination covered by pay-it-forward.

*Secondary analyses*

In addition to the pre-specified comparisons above, a decision-tree model will be built toanalyze the within-trial costs outcomes of pay-it-forward in comparison to

standard-of-care and free vaccination. We will use a healthcare provider perspective and the time horizon of the trial. A micro-costing approach will be used for the cost collection,including the start-up costs related to the pay-it-forward arm. We will examine factors associated with vaccine uptake in the standard-of-care arm and intervention arms. We will also determine factors associated with vaccine confidence and vaccine hesitancy inthe standard-of-care group and overall.

*Risk assessment and management*

According to the World Health Organization reports, influenza vaccines are well tolerated,and side effects are mild and transient in general. Common adverse events include soreness, redness, and/or swelling at the vaccination site, fever, muscle pain, headache, nausea which do not require medical attention.[12] Severe threatening allergic reaction after vaccination, such as difficulty breathing, hoarseness or wheezing, swelling around the eyes or lips, hives, paleness, weakness, or a fast heart beat or dizziness, are rare. These signs most likely happen within a few minutes to a few hours after the vaccine is given.

Participants who receive a vaccine will be required to stay at least half an hour in the vaccination clinic to be closely monitored by a designated nurse. They will be informed to self-monitor signs mentioned above for a few more hours after they leave the clinic. The contact number to a designated physician from the selected clinic will be provided toparticipants as emergency contact in the event of any severe adverse effects within 24 hours after the vaccination.

*Withdrawal of participants*

Participants can withdraw from the study at any time without giving any explanations. Withdrawal from the study would not affect the participant's medical care or other preventive services at the clinic in any way.

*Reporting of adverse events*

The local CDCs and vaccination clinics have established a strong surveillance system to monitor any adverse events associated with vaccinating patients. Each patient who receives an influenza vaccine is recorded in the clinical health information system and uploaded to the local CDCs' surveillance system. Patients with any adverse events will bedealt with by local health staff on a timely manner. The research team will leverage their existing platform for reporting any adverse events and keep a tracking record of adverse events. The research team will periodically meet with the designated health workers to make sure adverse events are properly treated.

*Data management*

Data will be managed according to the principles in accordance with good research practice specified by LSHTM Research Data Management Policy.[13] All data will be keptconfidential and accessible only to trained study staff. All consent forms, administrative and survey data will be transformed into digital format and data in non-digital formats will be stored in a securely locked cabinet at local institutes (Guangdong Second Provincial General Hospital in Guangzhou). Paper questionnaires will be filed and storedfor at least 10 years after project completion as per LSHTM's Records Retention and Disposal Schedule. Study databases will be managed by a dedicated data manager (DM).All data will be checked and cleaned before being updated to the main database. The databases will be regularly backed up on the local server where back-ups are maintained in the disaster recovery room. Copies of the databases will be transferred to LSHTM using the fully encrypted (SSL) server.

The main risks to data security relate to loss of electronic data and breach of participant confidentiality. With the acquisition of a dedicated office, all data on computers will be stored in this office which will be locked overnight. Only data entry personnel and study personnel will have access to any of the electronic data. After all checks have been completed, data will be entered into databases using desktops. The data manager will upload double entered data at the end of each day onto their computer. The Data Managerwill create weekly back-ups on an external password protected hard drive. When necessary or required, and copies of this data will be securely transferred to LSHTM using encrypted files. The risks for confidential breaching are minimal since most data will use anonymized IDs unique to each participant. We will ensure that all copies of the quantitative data files are password protected, with access to the password restricted to staff who need to work with the data. Also, we will report study results only at the aggregate level. To further minimize the risk of breaching confidentiality, all study staff will be trained and certified as part of MENISCUS-1 using the NIH online training course in Protecting Human Subject Research Participants and will be required to sign a staff confidentiality agreement form.

*Audits and inspections*

The study may be subject to audit by the London School of Hygiene & Tropical Medicine under their remit as sponsor, the Study Coordination Centre and other regulatory bodiesto ensure adherence to GCP.

*Insurance provision*

Trial insurance is covered by the trial sponsor.

**DSMB**

Given the low risks associated with a quasi-experimental study to improve existing publichealth service uptake, we believe having the project team as well as designate a data manager to meet periodically and monitor the data safety would suffice. DSMB will not be assembled in this particular project.

Research Proposal Version No.: 3

Research Proposal Version Date: Feb 9, 2021

*Trial registration*

This study was initially registered on Chinese clinical trials in September 2020. The proposal was last updated on 9 February 2021.

*Ethics*

We have obtained ethical approvals from Zhuhai CDC and the LSHTM ethics committee.

*Confidentiality*

Each participant will be assigned a unique ID and personal identifiable information will not be collected. Participant survey data will only be used for research purpose of this particular project. Participant survey data will be transformed into digital format and copies of the quantitative data files will be password protected. Only designated project staff will have access to this encrypted information. We will report study results only at the aggregate level which will be difficult for others to identify individual participants. Without participant permission, project team researchers will not share the information to a third party.

4. Hospital BJCs. Vaccination fraud alarmed the central government! In addition

to anger, you should also understand these. CN-Healthcare. 2018.

5. Kala W. Pregnant women are facing dilemma: both priority and contraindication for influenza vaccine. 2019 (accessed 22 Jan 2020).

11. Xueqiao W, Tom H. China pharma crackdown leads to flu vaccine shortage.

2018. https://www.ft.com/content/6829cd0e-f07b-11e8-ae55-df4bf40f9d0d (accessed 19 Jan 2020).

12. WHO. Information sheet: Observed rate of vaccine reactions - Influenza vaccine. . 2012. https://www.who.int/vaccine_safety/initiative/tools/Influenza_Vaccine_rates_information_sheet.pdf (accessed 19 Jan 2020).

13. LSHTM. Research data management policy. 2019. https://www.lshtm.ac.uk/sites/default/files/research_data_management_policy.pdf (accessed 19 Jan 2020).

Research Proposal Version No.: 3

Research Proposal Version Date: Feb 9, 2021

Supplementary file 1: Pay-it-forward leaflet

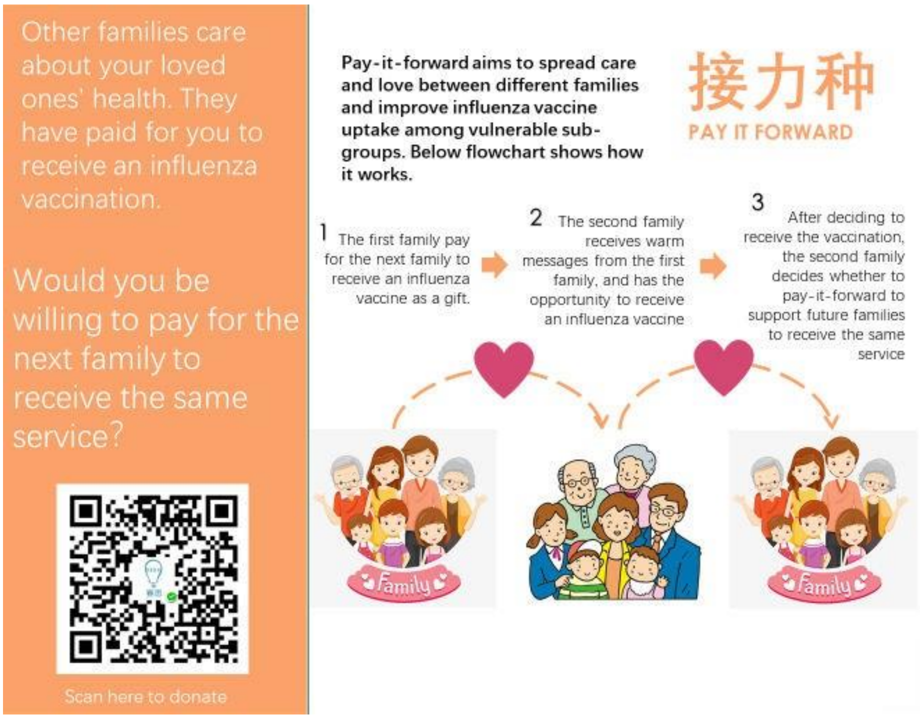

Research Proposal Version No.: 3

Research Proposal Version Date: Feb 9, 2021

Supplementary file 2: Examples of hand-written messages

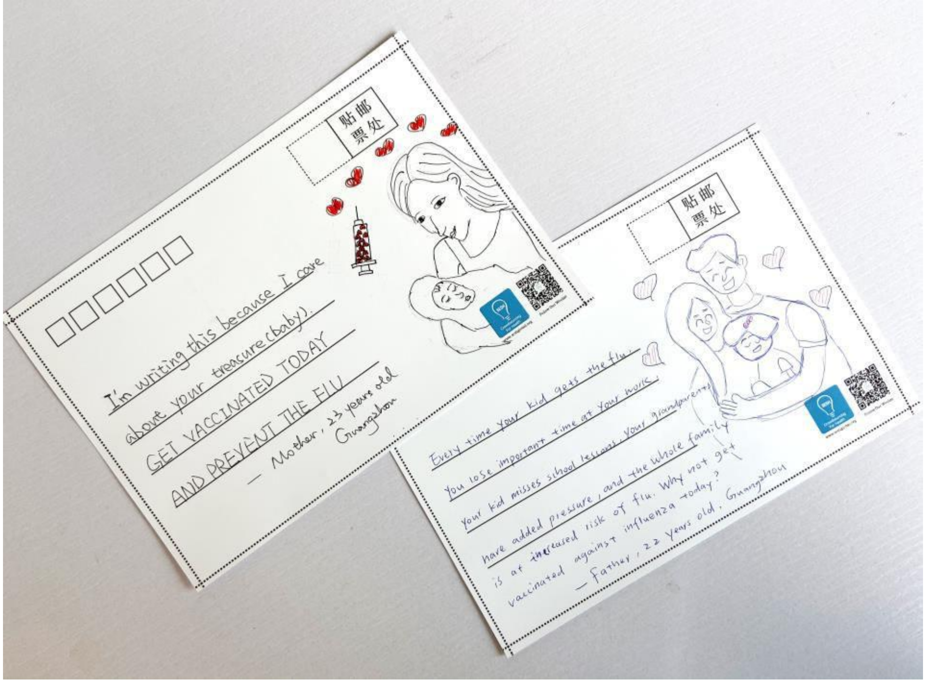
